# Ex vivo human leukemia blood model illustrates limitations of cancer-targeting PEGylated nanoparticles

**DOI:** 10.1101/2024.05.29.24308091

**Authors:** Yi Ju, Shiyao Li, Abigail Er Qi Tan, Emily H. Pilkington, Paul T. Brannon, Magdalena Plebanski, Jiwei Cui, Frank Caruso, Kristofer J. Thurecht, Constantine Tam, Stephen J. Kent

**Affiliations:** School of Science, RMIT University, Melbourne, Victoria 3000, Australia; Department of Microbiology and Immunology, Peter Doherty Institute for Infection and Immunity, The University of Melbourne, Melbourne, Victoria 3000, Australia; Materials Characterisation and Fabrication Platform, The University of Melbourne, Parkville, Victoria 3010, Australia; School of Health and Biomedical Sciences, RMIT University, Bundoora, Victoria 3083, Australia; Key Laboratory of Colloid and Interface Chemistry of the Ministry of Education, School of Chemistry and Chemical Engineering, Shandong University, Jinan, Shandong 250100, China; Department of Chemical Engineering, The University of Melbourne, Melbourne, Victoria 3000, Australia; Australian Institute for Bioengineering and Nanotechnology, University of Queensland, St Lucia 4072, Australia; Department of Clinical Haematology, The Royal Melbourne Hospital and Peter MacCallum Cancer Centre, Melbourne, Victoria, 3000, Australia; Faculty of Medicine, Dentistry and Health Sciences, University of Melbourne, Melbourne, Victoria, 3000, Australia; Melbourne Sexual Health Centre and Department of Infectious Diseases, Alfred Hospital and Central Clinical School, Monash University, Melbourne, Victoria 3000, Australia

## Abstract

Antibody-directed targeting of chemotherapeutic nanomaterials to primary human cancers could improve efficacy and reduce off-target toxicities. We developed an ex vivo model to study the targeting of primary human Chronic Lymphocytic Leukemia (CLL) in whole blood samples from 15 subjects with CLL. Anti-CD20 targeted polyethylene glycol (PEG)-based nanoparticles had generally efficient targeting of CLL cells and low off-target phagocytosis by neutrophils and monocytes. There was however substantial patient-patient variability (up to 164-fold difference in CLL targeting), driven in part by variance in pre-existing anti-PEG antibodies which reduced targeting effects. This suggests patients with lower PEG antibody levels may benefit more from targeted therapies. This was further exemplified by antibody-functionalized doxorubicin-containing PEGylated liposomes, which had relatively poor targeting of CLL in blood and high off-target uptake (significantly correlated with anti-PEG IgG levels in blood) and killing of almost all monocytes within 24 hours. Personalized low-fouling and non-PEGylated particle systems are needed to realize the potential of targeting chemotherapies. Overall, our human ex vivo model of tumor targeting by antibody-directed nanoparticles delineates limitations and opportunities of tumor-targeting nanomedicines.

---

Specific tumor targeting by nanomaterials to increase efficacy of anti-cancer therapies and reduce off-target effects is a long-held goal. A recurring problem for nanomaterials is their off-target uptake, clearance and cytotoxicity of phagocytes such as circulating neutrophils and monocytes.^1^ Immune suppression is the major dose-limiting and life-threatening toxicity of chemotherapeutics.^2^ A challenge for cancer treatments is providing a more favorable balance between anti-tumor activity and off-target toxicities.

Efficient targeting of nanomaterials to human cancers is yet to be realized and improved models of targeting nanomaterials to human cancers are needed towards this goal.^3^ Recent work has shown high targeting efficiency of immortalized leukemic cell lines in vitro and efficacy in leukemic cell line-engrafted immunodeficient mice.^4^ However, experiments studying *in vitro* targeting of tumor cell lines do not capture the complexities of targeting primary human cancers in the presence of autologous bystander cells. Further, in-bred mouse models typically have restricted immune systems developed in pathogen-free environments and rely on induction or transplantation of tumors on a highly immunodeficient background that is unlikely to reliably model human malignancy. Better human models are needed.

Antibodies have exquisite specificity and are commonly used to target nanomedicines. However, a major problem with antibody-targeted nanoparticles is that the large surface antibody molecule leads to increased “fouling” *in vivo* or *ex vivo* through the generation of enhanced biomolecular coronas and consequent uptake by phagocytes.^5,6^ This inadvertently reduces or ameliorates the effect of the targeting. Indeed, Sivaram *et al* found that overly high levels of a solid tumor-targeting antibody on the surface of nanoparticles led to increased uptake of the nanoparticles by fresh human blood phagocytes ex vivo and reduced targeting in mouse models.^7^ The balance between tumor targeting and off-target effects has however not to our knowledge been probed in primary human cancer models in the presence of both the tumor target and off-target immune cells.

We previously pioneered whole human blood cell models to study the association of nanoparticles and human immune cells.^8^ The complex mix of human blood mimics the environment that particles would interact with if administered intravenously. We previously observed marked variability across healthy human donors in the capacity of their plasma to mediate fouling of nanomaterials and increased uptake by phagocytic cells.^9^ For the many nanomaterials that contain poly(ethylene)-glycol (PEG), pre-existing PEG antibodies mediate much of the variability in immune cell uptake observed.^10^ High levels of PEG antibodies in blood result in increased fouling of nanoparticles and high levels of uptake by phagocytes such as neutrophils and monocytes.

We reasoned that spontaneous human blood leukemias provide a milieu in which the blood cell tumor co-circulates with other blood cell components including phagocytes such as neutrophils and monocytes. This provides an *ex vivo*, fully human, outbred model in which to assess the balancing of targeting tumors with nanoparticles versus the removal of nanoparticles by phagocytes. Herein we recruited subjects with Chronic Lymphocytic Leukemia (CLL), a cancer with an abundance of malignant CD20+ B cells in blood. CLL is a chronic malignancy with suppressed levels of normal B cells but relatively preserved numbers and functions of most other blood immune cells. Fresh blood from subjects with CLL was employed to study the ability of antibody-targeted nanoparticles to simultaneously target the B cell tumor and avoid off-target uptake of particles by other blood immune cells, particularly granulocytes and monocytes.

## Results

### Establishing targeted nanoparticles and a B and T cell tumor cell model in human blood

To advance a better understanding of targeting blood cell cancers, we first established a range of targeted PEG-based nanomaterials and evaluated their activity when incubated with fresh human donor blood which had B and T cell immortalized cell lines spiked into. We previously developed 3 particle models spanning a range of higher, medium or lower-fouling materials.^9^ That is, PEGylated mesoporous silica (PEG-MS) particles are high-fouling, PEGylated liposomes containing doxorubicin (that were formulated with the same lipid composition and drug/lipid ratio as FDA-approved liposomal doxorubicin agent Doxil) are medium fouling, and pure PEG particles are lower fouling (Fig 1a). We employed bispecific antibodies (BsAb) to functionalize the nanoparticles, with one antibody arm binding to the PEG on the nanoparticle and the other binding to either the B cell marker CD20 or the T cell marker CD28. We used T cell targeting as a control for the B cell targeting in addition to untargeted particles. We established that all 3 particle systems efficiently and specifically target the relevant cell lines and primary T and B cells *in vitro* (Supplementary Fig. 1 and 2). The three types of particles were fully characterized with similar size and polydispersity before and after BsAb functionalization (Supplementary Fig. 3 and Supplementary Table 1). After a one-hour co-culture, we used flow cytometry (gating strategy shown in Supplementary Fig. 4,5) to identify whether the fluorescent targeted nanoparticles were binding to the B or T cell line targets and/or binding to off-target cells in blood such as monocytes and granulocytes (Fig. 1b).

**Table 1.** Demographic information, cell count, CD20/CD28 expression data of 15 CLL patients.

| Patient ID | Age | Sex | Total WBC ($\times 10^9/L$) | CLL ($\times 10^9/L$) | CLL (%) in WBC | CD20+ (%) in CLL | T cell (%) in WBC | CD28+ (%) in T cell |
| --- | --- | --- | --- | --- | --- | --- | --- | --- |
| 1 | 86–90 | M | 29 | 19 | 65 | 98 | 6 | 59 |
| 2 | 56–60 | F | 45 | 31 | 69 | 99 | 8 | 84 |
| 3 | 66–70 | M | 41 | 32 | 80 | 99 | 1 | 77 |
| 4 | 46–50 | F | 49 | 35 | 71 | 99 | 9 | 87 |
| 5 | 66–70 | M | 9 | 1 | 7 | 100 | 27 | 82 |
| 6 | 76–80 | M | 42 | 33 | 78 | 91 | 2 | 87 |
| 7 | 71–75 | M | 24 | 14 | 56 | 100 | 9 | 74 |
| 8 | 76–80 | F | 50 | 36 | 71 | 86 | 1 | 96 |
| 9 | 61–65 | F | 66 | 48 | 73 | 90 | 4 | 85 |
| 10 | 51–55 | M | 32 | 23 | 73 | 96 | 3 | 95 |
| 11 | 56–60 | M | 18 | 11 | 64 | 95 | 11 | 84 |
| 12 | 71–75 | F | 20 | 11 | 54 | 100 | 12 | 62 |
| 13 | 66–70 | F | 9 | 3 | 32 | 80 | 11 | 91 |
| 14 | 51–55 | M | 9 | 6 | 68 | 100 | 14 | 79 |
| 15 | 71–75 | M | 65 | 50 | 77 | 96 | 2 | 82 |
| Mean (range) | 67 (49–90) | 60% M | 34 (9–66) | 24 (1–50) | 63% (7–80%) | 95% (80–100%) | 8% (1–27%) | 82% (59–96%) |

**Fig. 1.**
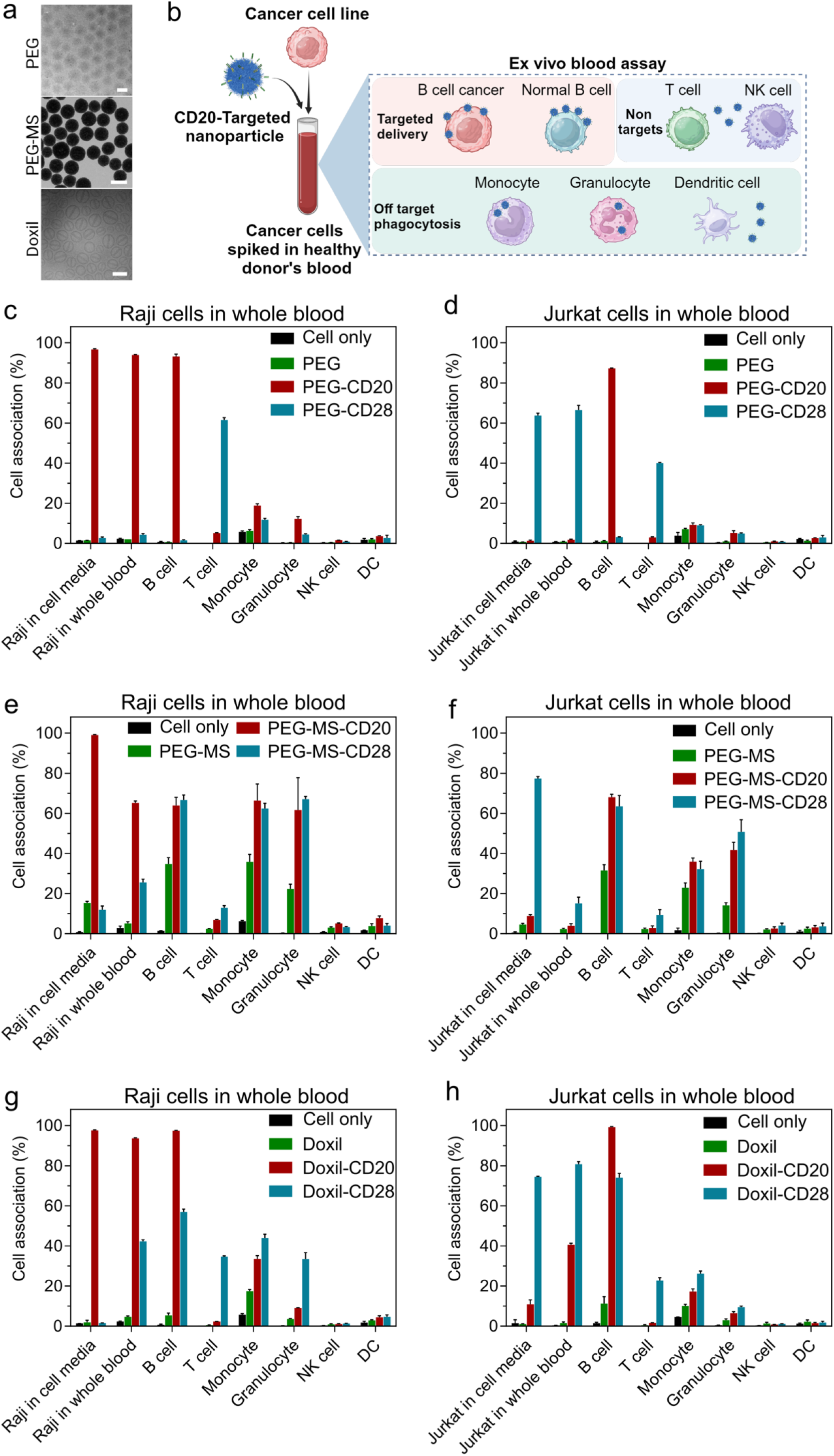
Evaluation of cancer cell targeting in whole human blood model. a) Transmission electron microscopy (TEM) images of PEG and PEG-MS particles and a cryogenic TEM image of Doxil nanoparticles. Scale bars are 500 nm (PEG and PEG-MS) and 100 nm (Doxil), respectively. b) Schematic illustration of the whole human blood model that contains both cancer cells and blood cells to assess nanoparticle targeting in comparison to conventional cell culture model. Created with BioRender.com. c-h) Cancer cell targeting in cell culture media versus in whole human blood of BsAb-functionalized PEG, PEG-MS and Doxil nanoparticles. Raji (b-d) or Jurkat (e-g) cells were added into cell culture media or fresh human blood from a healthy donor, followed by incubating with nanoparticles for 1 h at 37 °C and subsequent analysis by flow cytometry. Cell association (%) refers to the proportion of each cell type with fluorescence above background, stemming from fluorescence-labeled particles (See gating strategy in Supplementary Fig. 4, 5). Cell association (%) data are shown as the mean of three independent experiments (using the fresh blood from the same donor), with at least 400,000 leukocytes analyzed for each experimental condition studied. Cell only control groups represent the respective cell populations without particle incubation. NK: natural killer cells; DC: dendritic cells.

**Fig. 2.**
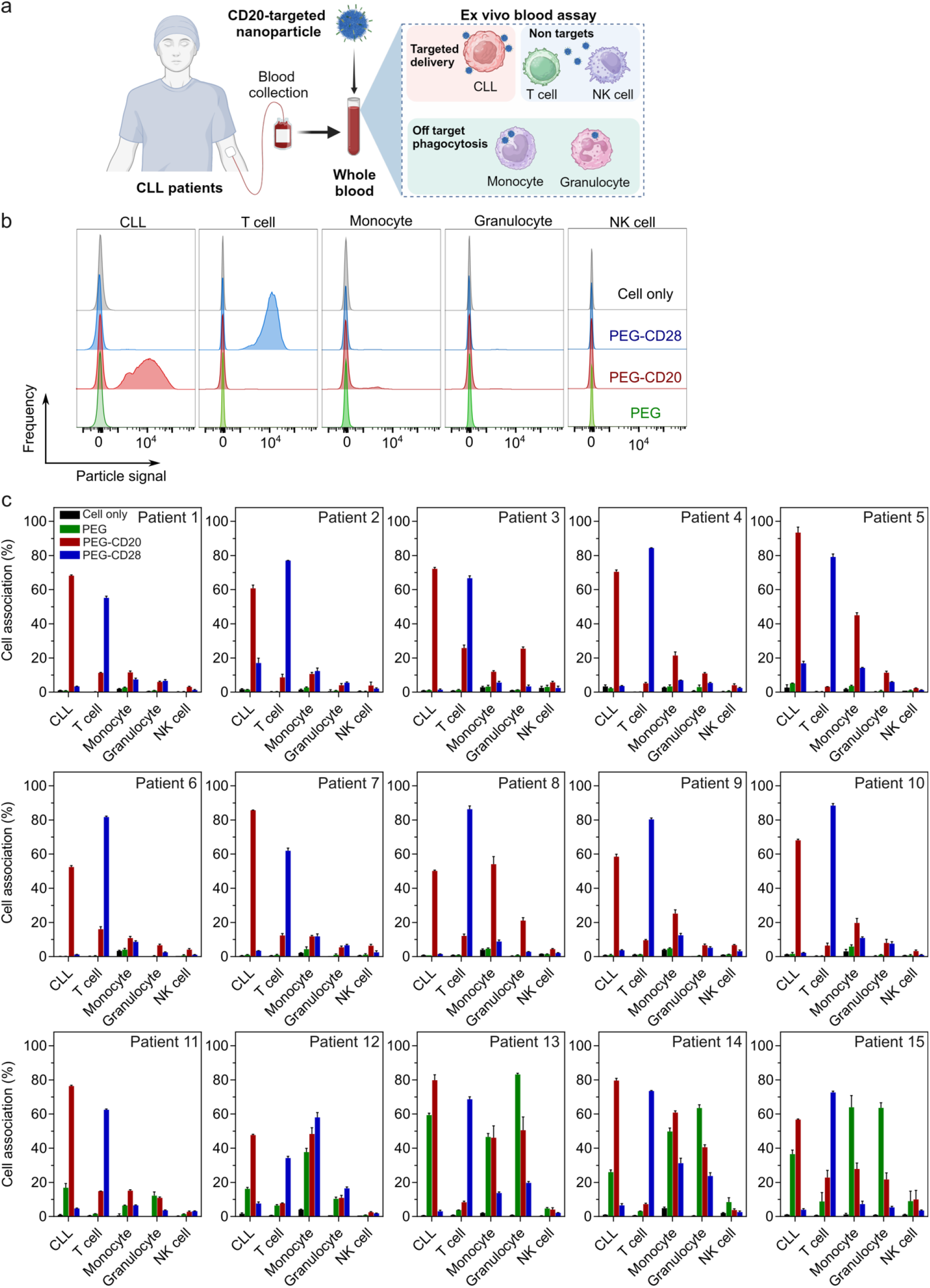
Cancer cell targeting of BsAb-functionalized PEG particles in the whole blood of CLL patients. a) Schematic illustration of PEG particles functionalized with anti-PEG/anti-CD20 BsAbs were incubated with fresh whole blood of a CLL patient for 1 h at 37 °C and subsequent analysis by flow cytometry. b) Flow cytometry histograms represent the cell association of PEG, PEG-CD20, and PEG-CD28 particles in the blood of a CLL patient. c) CLL targeting of BsAb-functionalized PEG particles in the whole blood of the 15 CLL patients (n=15, see Table 1). Cell association (%) refers to the proportion of each cell type with fluorescence above background, stemming from fluorescence-labeled particles (See gating strategy in Supplementary Fig. 8). Cell association (%) data are shown as the mean of three independent experiments (using the fresh blood from each donor), with at least 400,000 leukocytes analyzed for each experimental condition studied. Cell only control groups represent the respective cell populations without particle incubation. Donor IDs are arranged based on monocyte−PEG particle association (from low to high).

**Fig. 3.**
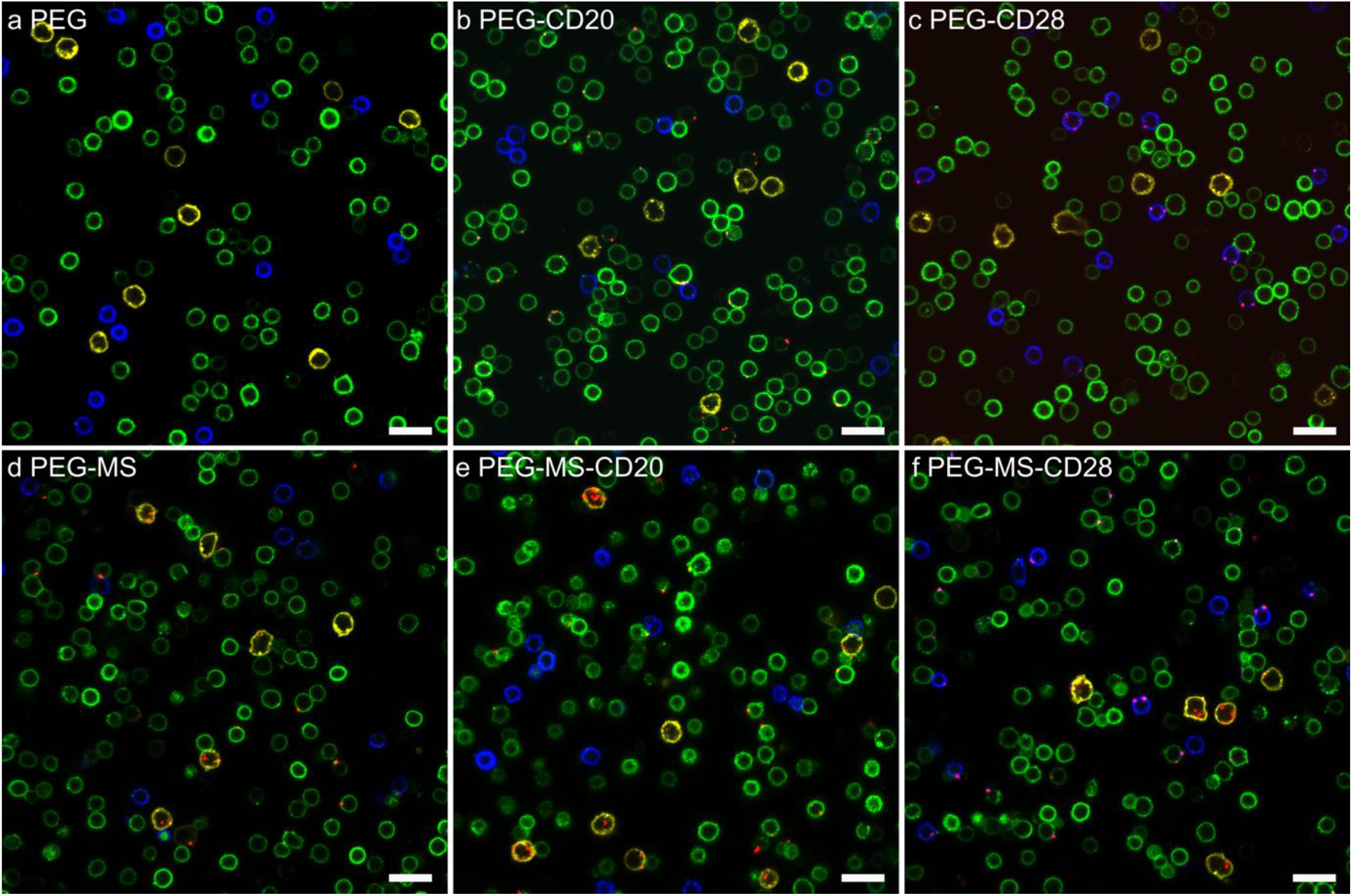
Confocal microscopy images of BsAb-functionalized PEG and PEG-MS particle association with blood cells from a CLL patient. a-c) PEG and d-e) PEG-MS particles functionalized with or without anti-PEG/anti-CD20 or anti-PEG/anti-CD28 BsAbs were incubated with peripheral blood mononuclear cells (PBMCs) from a CLL patient for 1 h at 37 °C, followed by phenotyping cells with antibody cocktails and imaged by confocal microscopy. Green: CD20+ CLL cells; blue: CD3+ T cells; yellow: CD14+ monocytes; red: fluorescence-labelled PEG or PEG-MS particles. Scale bar: 20 μm.

**Fig. 4.**
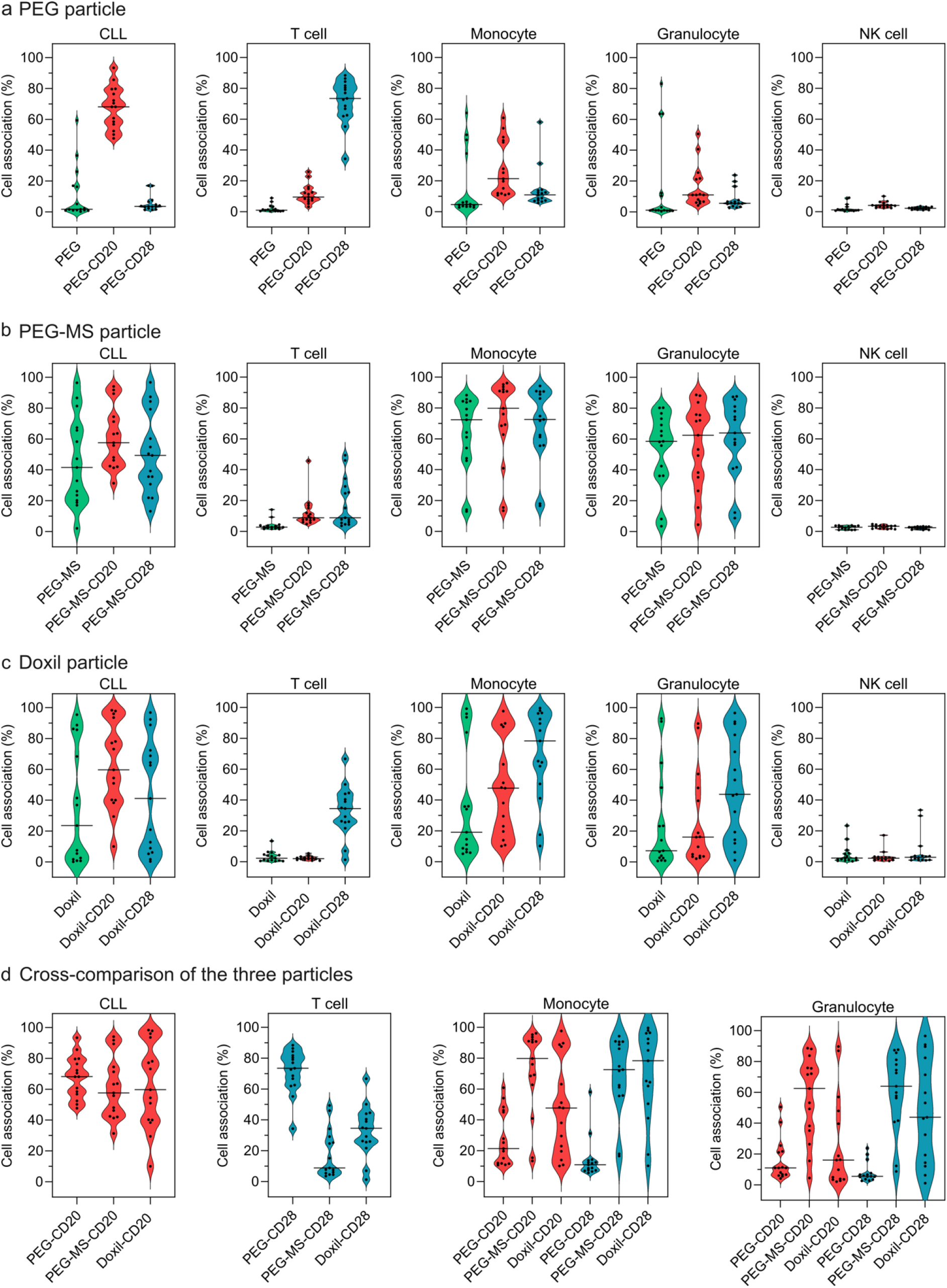
Cancer cell targeting of PEG, PEG-MS, and Doxil nanoparticles in the whole blood of 15 CLL patients. a-c) Violin plots summarizing the BsAb-functionalized PEG, PEG-MS, and Doxil nanoparticle association with CLL cells, T cells, monocytes, granulocytes, and NK cells in CLL patients’ blood after 1 h incubation at 37 °C. d) Cross-comparison of the association of the three particles with CLL cells, T cells, monocytes, and granulocytes in CLL patients’ blood. Each data point is the mean of three independent experiments (using the same batch of fresh blood from each donor). The median cell associations across 15 CLL patients are shown as solid lines in the violin plots.

**Fig. 5.**
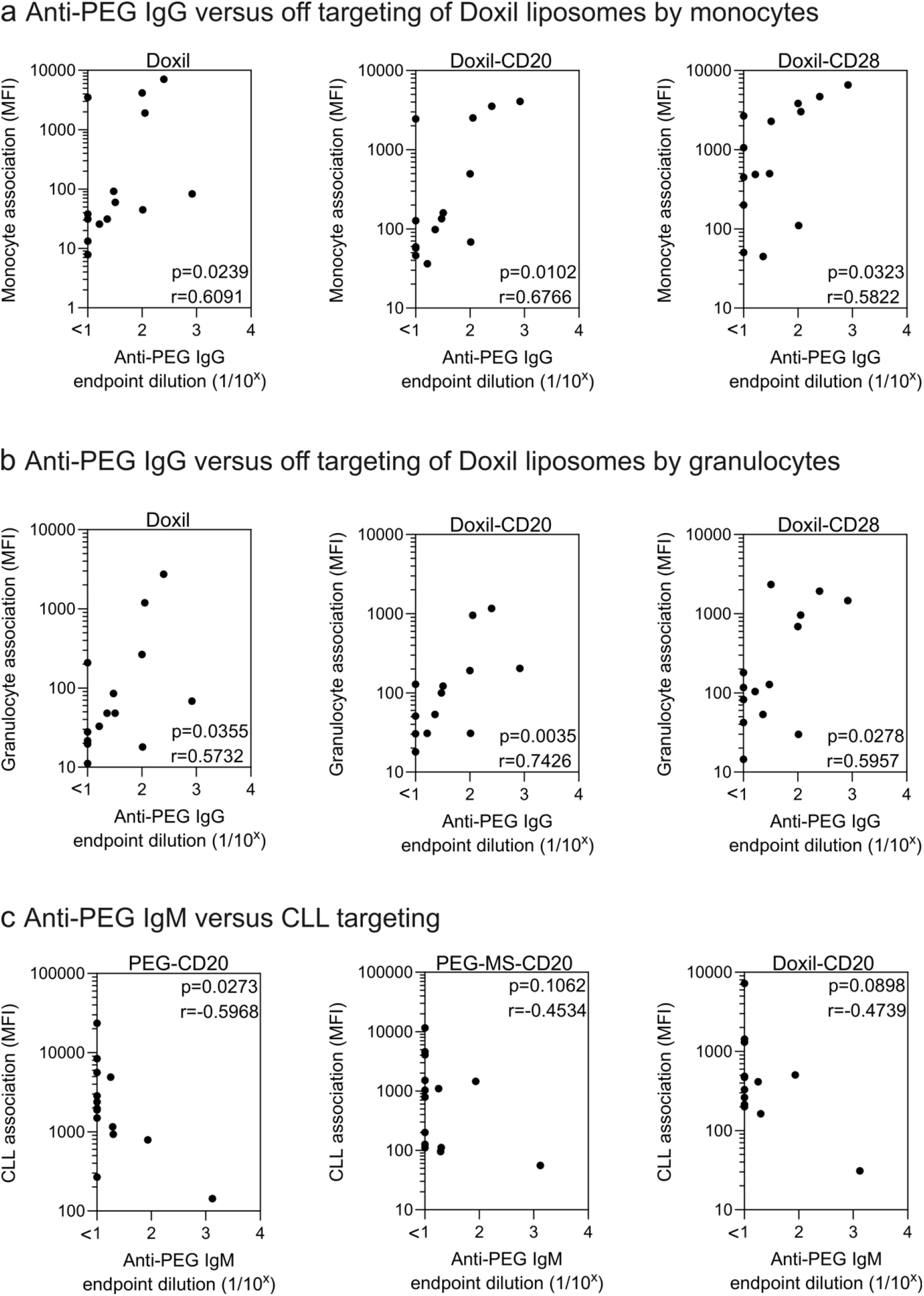
Anti-PEG antibodies influence the particle phagocytosis and CLL targeting in the blood of CLL patients. a,b) Plasma anti-PEG IgG levels positively correlate with the phagocytosis of Doxil, Doxil-CD20, and Doxil-CD28 by monocytes and granulocytes. c) Plasma anti-PEG IgM levels negatively correlate with CLL targeting of PEG-CD20 particles. Cell association (MFI) refers to the median fluorescence index of each cell type, stemming from fluorescence-labeled particles. Plasma anti-PEG IgG and IgM levels were determined by ELISA (Supplementary Fig. S10). Statistics were assessed by Spearman correlation analysis (n=14).

When the lower-fouling PEG particles with the anti-CD20 BsAb are incubated with Raji B cells spiked into whole blood there is efficient targeting to both the Raji B cell line and normal B cells but minimal off-target binding to phagocytes (Fig. 1c). Similarly, when PEG particles with the anti-CD28 BsAb are incubated with the Jurkat T cells spiked into whole blood there is efficient targeting to both the Jurkat T cell line and normal T cells but minimal binding to phagocytes (Fig. 1d). The intermediate fouling Doxil particles and high-fouling PEG-MS particles show less efficient targeting and increasing off-target uptake by phagocytes. The particles with the targeting BsAb are generally higher fouling than the untargeted particles (Fig. 1e-h), consistent with previous work showing particle surface antibodies can reduce stealthiness.^7,11^ These experiments established an in vitro model of a range of targeted nanomaterials to B and T cells in the presence of off-target blood cells that allowed us to recruit human participants for a clinically relevant cancer-targeting model.

### Targeting nanoparticles to the B cell tumor in the blood of CLL patients

Having set up a blood model of targeting nanoparticles to B cells, we proceeded to recruit 15 participants known to have CLL with significant numbers of malignant B cells in their blood. In most subjects, the malignant B cells out-numbered normal white cells in blood, as expected for CLL not in remission (Table 1, Supplementary Fig. 6). Almost all the malignant B cells expressed CD20. None or very few normal B cells (< 0.5% of total CD19+ B cell populations, Supplementary Fig. 7) were identified in the blood of CLL subjects and were thus not included in the targeting study. Normal T cells were present as expected and on average 82% expressed CD28, the target of control-target nanoparticles.

**Fig. 6.**
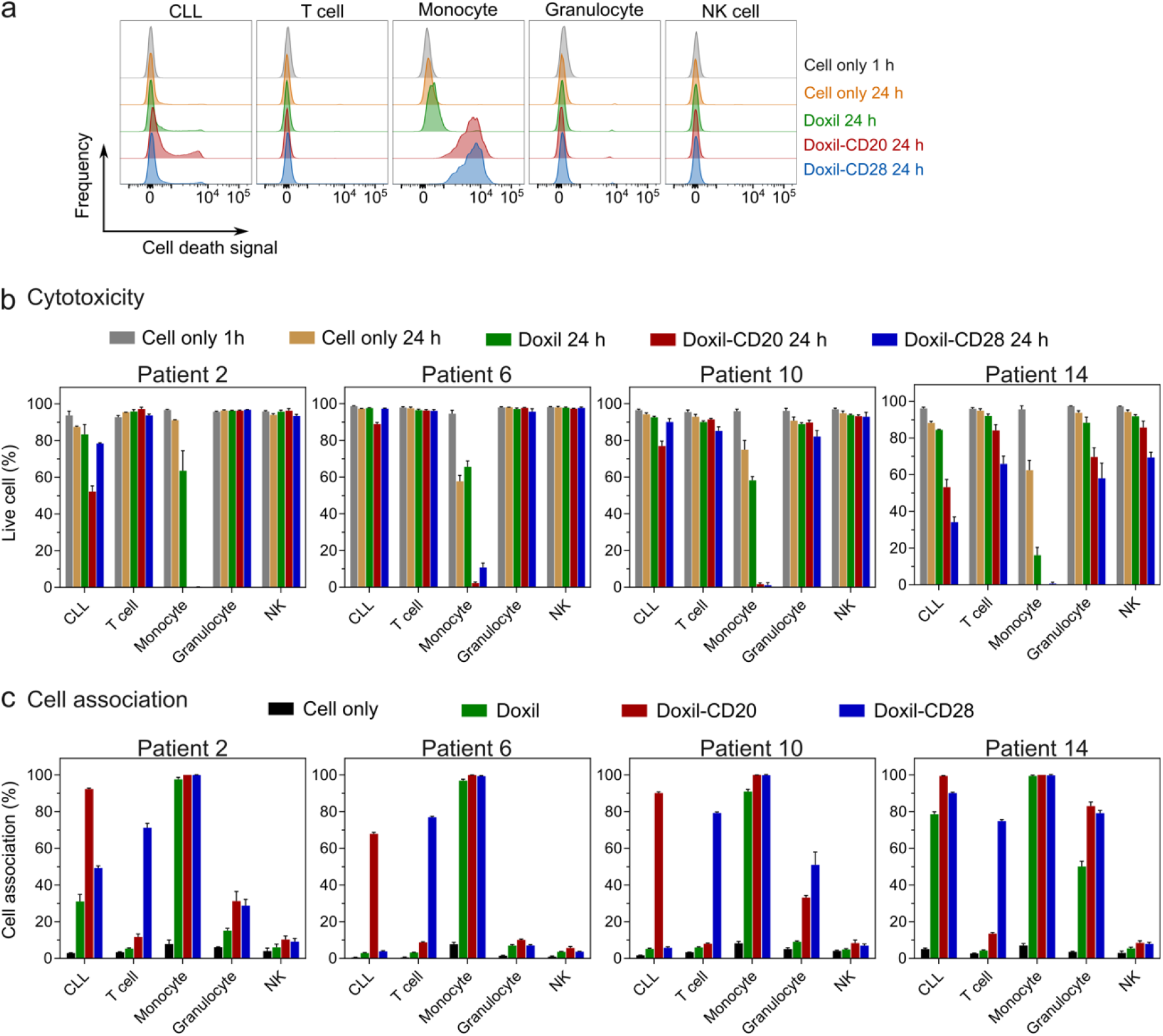
Targeted and off-targeted cytotoxicity of BsAb-functionalized Doxil nanoparticles to CLL and healthy immune cells in blood of CLL patients. a) Flow cytometry histograms represent the cell death of CLL cells, T cells, monocytes, granulocytes, and NK cells after treating with Doxil nanoparticles in the blood of a CLL patient (Patient 2). b) Cytotoxicity and c) cell association of BsAb-functionalized Doxil nanoparticles in the whole blood of four CLL patients. Doxil nanoparticles were included with the whole blood of CLL patients for 24 h at 37 °C, followed by phenotyping cells with antibody cocktails, labelling dead cells with SYTOX^™^ AADvanced^™^ Dead Cell Stain, and analysis by flow cytometry.

We incubated our range of 9 targeted nanoparticles with fresh blood for 1 hour from all 15 CLL participants (Fig. 2a), followed by analysis with flow cytometry (Fig. 2b, gating strategy shown in Supplementary Fig. 8). Results of the lower-fouling PEG particles are shown in Fig. 2c. For most subjects, there was efficient targeting of the CD20-targeted PEG particles to the CLL with much less off-target binding to T cells, monocytes or granulocytes. Similarly, the CD28-targeted PEG particles targeted T cells with generally minimal off-target binding to the malignant B cells, monocytes or granulocytes. We illustrated this visually by confocal microscopy in blood cells from one subject in Fig. 3b, where the B cell tumor (in green), but not T cells (in blue) or monocytes (in yellow) can commonly be seen to have CD20-targeted PEG particles on their surface. CD28-targeted PEG particles bind primarily to T cells only and untargeted PEG particles rarely bind to any cells (Fig. 3a,c).

It was notable that several subjects (Patient #11-15 in Fig. 2c) had much higher association of a range of the PEG particles by monocytes and granulocytes. Although targeting of CD20-functionalized PEG particles was generally maintained, there was substantial interpatient variability in CLL targeting (up to 164-fold difference, supplementary Fig. 9a) and off-targeting association of phagocytes (up to 6- and 12-fold difference in monocytes and granulocytes, respectively, Fig.4a) across all subjects.

In contrast to the PEG particles, the higher-fouling PEG-MS particles had generally poor targeting and high off-target binding across most donors. Results summarized across the 15 subjects are shown in Fig. 4b and supplementary Fig. 9b with the primary data for each subject shown in Supplementary Fig. 10. The PEG-MS particles had minimal specific CLL targeting with most monocytes and granulocytes associating with PEG-MS particles. This is illustrated visually in Fig 3e,f, where monocytes (in yellow) commonly have phagocytosed many particles, with rarer specific binding to the CLL or T cell targets. This illustrates how particle chemistry across the 3 nanomaterials studies had a major effect on targeting and off-target effects.

Doxil is a clinically used liposome chemotherapeutic nanoparticle.^12^ The targeted Doxil nanoparticles showed an intermediate phenotype to that of the PEG and PEG-MS particles. Generally, modest targeting of the CD20-targeted Doxil liposomes to the CLL cells was observed and substantial off-target uptake by monocytes and granulocytes was observed (Fig. 4c, Supplementary Fig. 9c, 11). The 3 particle systems are compared in Fig. 4d to illustrate this clearly. The large range of targeting and off-target effects across the 15 donors is notable across all 3 particle systems.

### PEG antibodies are associated with higher off-target phagocyte uptake

The large range of off-target binding of the PEGylated nanoparticles across the 15 CLL donors was similar to our previous studies in 23 healthy blood samples where we also observed broad interpersonal differences in phagocyte-nanoparticle uptake.^9^ This previous work linked interpersonal differences to differences in the protein corona forming on the particles, particularly the presence of immunoglobulin molecules. We recently developed methods to analyze PEG-specific antibodies and found that higher levels of PEG-specific antibodies induced by SARS-CoV-2 mRNA vaccines in humans were linked to higher uptake of PEGylated nanoparticles by blood phagocytes.^10^ This raised the hypothesis that the range off-target binding of targeted nanomaterials could be linked at least in part to PEG-specific antibodies binding to and fouling the PEGylated nanoparticles.

We measured PEG-specific IgG and IgM antibodies across 14 of the CLL blood donors and found a large interpatient variance (up to 83- and 133-fold difference in titers of IgG and IgM, respectively, supplementary Fig. 12). The association of PEGylated Doxil liposomes with both granulocytes and monocytes significantly correlated with PEG-specific IgG (Fig. 5a,b). This was true for both untargeted and BsAb targeted Doxil nanoparticles as expected since all 3 have surface PEG. We did not observe significant correlations with PEG antibodies and uptake of PEG or PEG-MS nanoparticles by phagocytes (Supplementary Fig. 13, 14), perhaps owing to the overall very low or very high fouling of PEG and PEG-MS particles respectively, reducing the power to observe additional contributions of PEG antibodies.

We also studied the influence of PEG-specific antibodies on the ability of the CD20-functionalized particles to target CLL cells within whole blood. We hypothesized that higher PEG antibodies could either reduce particle availability through off-target binding or reduce targeting due to PEG antibodies contributing to fouling and/or target antibody masking. We found significantly reduced targeting of CLL by CD20-targeted PEG particles in the presence of higher levels of PEG-specific IgM, with trends observed for CD20-targeted PEG-MS and Doxil particles (Fig. 5c, Supplementary Fig. 15b). Interestingly, we did not observe reduced targeting with higher PEG-specific IgG (Supplementary Fig. 15a), suggesting the larger PEG-specific pentavalent IgM molecule binding to may have a more important role in occluding or sterically hindering the targeting BsAb molecule.

### Cytotoxicity effects of targeted Doxil particles on both CLL and phagocytes

A goal of chemotherapy is to kill the tumor while minimizing off-target killing of healthy immune cells. We further advanced our whole blood CLL model to study this *ex vivo* across 4 patients by using a longer (24hr) incubation to allow for time to observe cell death. We simultaneously studied both killing and targeting of CLL and other blood cells through employing a live-dead stain and flow-cytometry after 24h incubation of Doxil particles (which contain the chemotherapeutic agent doxorubicin) with whole blood (Fig. 6a). In all 4 subjects, we observed near complete death of monocytes by the CD20 or CD28 targeted Doxil, with a more limited effect by the untargeted Doxil (Fig. 6b). The death of monocytes was consistent with their high uptake of Doxil liposomes and their sensitivity to the doxorubicin (Fig. 6c). There was only modest targeted killing of the CLL cells over the 24h incubation by the CD20-targeted Doxil (9−31% killing over untargeted Doxil). This likely reflects both the modest targeting, limited intracellular uptake of the liposomes and the relative resistance of the CLL cells to doxorubicin. One subject (patient 14) had no specific targeting – this subject had high uptake of all particles by phagocytes and incidentally had high levels of PEG IgG. Overall, this data is consistent with a model where phagocytes are commonly killed through off-target uptake by higher-fouling nanoparticles, potentially abetted through opsonization by PEG-specific antibodies.

## Discussion

The effectiveness of chemotherapy is limited in large part by off-target toxicities on the healthy immune system. We developed a fully human ex vivo model of targeting a primary human B cell malignancy (CLL) in the presence of immune cells within blood. This model allowed us to identify important issues around targeting human cancers with antibody-targeted nanomaterials.

We observed a remarkably large person-to-person variability in the capacity of nanomaterials to target primary human cancer in blood and avoid off-target uptake and killing of phagocytes. We identified that some of this variability is due to anti-PEG antibodies opsonizing PEGylated nanomaterials. Anti-PEG antibodies will bind to the surface of PEGylated nanoparticles, leading to complement deposition of complement reaction products on nanoparticles.^13^ This will result in the formation of an anti-PEG antibody-modulated protein corona around the nanoparticle. The protein corona will impede the efficiency of the hydrophilic properties of PEG to reduce the fouling and off-target uptake of the PEGylated materials by phagocytes. We previously identified antibody molecules and complement as major components of the corona modulating interactions of PEGylated nanomaterials with circulating phagocytes.^9^ Further, the PEG antibodies and the associated protein corona bound to antibody-coated nanoparticle will likely have an effect of sterically hindering the ability of a targeting antibody to engage its ligand, reducing targeting efficiency, a phenomenon that has been observed in other types of PEGylated targeted nanoparticles.^14^

In this study we found subjects with CLL and low PEG antibody levels had higher targeting efficiency of antibody-functionalized PEGylated nanomaterials and lower off-target phagocyte association. Our results predict subjects with low PEG antibodies would respond more effectively and safely to targeting nanotherapeutics that containing PEG, a testable hypothesis for future clinical trials or in retrospective analyses. For subjects with higher PEG antibodies, improved low-fouling nanomaterials not containing PEG are needed, a field of considerable interest.^15-19^ The widespread use of PEG in cosmetics and other materials,^20,21^ as well as the incorporation of PEG in current SARS-CoV-2 mRNA vaccines,^22^ may mean that more people will have higher PEG antibodies and respond more poorly to PEGylated cancer nanomaterials in the future.

We also observed distinct patterns across the range of 3 nanoparticle systems studied that have implications for future cancer nanotherapeutics. We found that lower-fouling nanoparticles, such as the pure PEG particles we studied,^23,24^ were critical to achieve high targeting and avoid off-target uptake in most subjects. Even in the absence of high levels of plasma PEG antibodies, CD20 targeting PEG-MS and Doxil systems rarely had both efficient targeting and low off-target effects. This work is consistent with the overall high toxicity of Doxil liposomes observed clinically.^25^ Monocytes avidly phagocytosed targeted or untargeted Doxil liposomes and were completely killed within 24hrs of exposure ex vivo. Less than half of the CLL cells were specifically killed by CD20-targeted Doxil liposomes within a similar time frame. Although sobering, these results provide an *ex vivo* benchmark upon which to improve targeting and reduce off-target effects of novel chemotherapeutic nanomaterials. Each *ex vivo* test of both targeting and off-target effects requires only 100μl of blood, providing a system with reasonable throughput for the rational selection of improved cancer nanotherapeutics.

In conclusion, we provide a novel ex vivo blood model that allows simultaneous study of cancer targeting and cancer toxicity by nanomaterials. We found that nanoparticle fouling levels and anti-PEG antibodies were major determinants of successful cancer targeting and reduced off-target effects on healthy immune cells. This work should promote and guide the generation of both improved cancer nanomaterials and the improved selection of patients for personalized treatments.

## Methods

### Ethics statement

The study protocols were approved by the University of Melbourne and Royal Melbourne Hospital Human Research Ethics Committee (# 2057981.1, 13/36, 22993-34586-3) and all associated procedures were carried out in accordance with the approved guidelines.

### CLL Patient recruitment and sample collection

CLL patients were recruited through contacts with the investigators and invited to provide blood samples. For all participants, whole blood was collected with sodium heparin anticoagulant. Plasma was collected and stored at −80 °C, and PBMCs were isolated via Ficoll-Paque separation, cryopreserved in 10% DMSO/FCS and stored in liquid nitrogen.

### Whole human blood model spiked with B or T cell lines

The fresh human blood was collected from a healthy volunteer into sodium heparin vacuettes (Greiner Bio-One) and counted with a CELL-DYN Emerald analyzer. Raji B cells and Jurkat T cells were labelled with CellTrace™ Yellow Cell Proliferation Kit at 1.5 μM in PBS at 37 °C for 20 mins, and subsequently diluted and incubated with completed RPMI-1640 media for 5 mins at 37 °C, followed by washing with completed RPMI-1640 media twice. Pre-labelled Raji or Jurkat cells (1 × 10^5^) were spiked into 100 μL fresh human blood, followed by adding fluorescence-labelled PEG (2 × 10^7^), PEG-MS (2 × 10^7^) and Doxil nanoparticles (0.44 μg based on lipid) with or without BsAb functionalization. After 1 h incubation at 37 °C, the whole blood was incubated on ice for 10 mins, and the red blood cells were lysed using Pharm Lyse buffer (4 mL) and removed by centrifugation (500*g*, 5 min). After being washed twice with FACS buffer, the cells were phenotyped at 4 °C for 1 h using an antibody cocktail consisting of CD3 AF700 (SP34-2, BD), CD14 APC-H7 (MΦP9, BD), CD66b BV421 (G10F5, BD), CD45 V500 (HI30, BD), CD56 PE-Cy7 (HCD56, BioLegend), lineage-1 (Lin-1) cocktail FITC (BD), HLA-DR PerCP-Cy5.5 (G46-6, BD), and CD19 BV650 (HIB19, BioLegend) in titrated concentrations. To assess the CD20 and CD28 expression, CD20 AF647 (2H7, BioLegend) or CD28 AF647 (CD28.2, BioLegend) were incubated with cells at 4 °C for 1 h. To assess the CD20 and CD28 expression, CD20 AF647 (2H7, BioLegend) or CD28 AF647 (CD28.2, BioLegend) were incubated with cells at 4 °C for 1 h. The cells were subsequently washed twice with FACS wash buffer (4 mL, 500*g*, 7 min) to remove unbound antibodies and fixed with 4% paraformaldehyde in PBS and directly analyzed by flow cytometry (LSRFortessa, BD Bioscience). The data were processed using FlowJo V10. Cell association (%) refers to the proportion of each cell type with fluorescence above background, stemming from fluorescence-labeled particles (See gating strategy in Supplementary Fig. 4,5). Cell association (%) data are shown as the mean of three independent experiments (using the fresh blood from each donor), with at least 400,000 leukocytes analyzed for each experimental condition studied.

### Whole blood model from CLL patients

The fresh human blood was collected from CLL patients into sodium heparin vacuettes (Greiner Bio-One) and counted with a CELL-DYN Emerald analyzer. The fresh blood (100 μL) was subsequently incubated with fluorescence-labelled PEG (2 × 10^7^), PEG-MS (2 × 10^7^), and Doxil nanoparticles (0.44 μg based on lipid) with or without BsAb functionalization at 37 °C for 1 h. After incubation, the whole blood was incubated on ice for 10 mins, and the red blood cells were lysed using Pharm Lyse buffer (4 mL) and removed by centrifugation (500*g*, 5 min). After being washed twice with FACS buffer, the cells were phenotyped at 4 °C for 1 h using an antibody cocktail consisting of CD3 AF700 (SP34-2, BD), CD14 APC-H7 (MΦP9, BD), CD66b BV421 (G10F5, BD), CD56 PE-Cy7 (HCD56, BioLegend), CD19 BV650 (HIB19, BioLegend), CD5 BV786 (UCHT2, BD) and CD22 PE (S-HCL-1, BD) in titrated concentrations. To assess the CD20 and CD28 expression, CD20 AF647 (2H7, BioLegend) or CD28 AF647 (CD28.2, BioLegend) were incubated with cells at 4 °C for 1 h. The cells were subsequently washed twice with FACS wash buffer (4 mL, 500*g*, 7 min) to remove unbound antibodies and fixed with 4% paraformaldehyde in PBS and directly analyzed by flow cytometry (LSRFortessa, BD Bioscience). The data were processed using FlowJo V10. Cell association (%) refers to the proportion of each cell type with fluorescence above background, stemming from fluorescence-labeled particles (See gating strategy in Supplementary Fig. 8). Cell association (%) data are shown as the mean of three independent experiments (using the fresh blood from each donor), with at least 400,000 leukocytes analyzed for each experimental condition studied.

### Confocal microscopy images of CLL PBMCs

Frozen PBMCs from a CLL patient were thawed, washed with RPMI 1640 (serum-free) media twice, and counted with a CELL-DYN Emerald analyzer. The washed PBMCs (3 × 10^6^) were dispersed in 100 μL RPMI 1640 (serum-free) media, followed by adding 100 μL human plasma from the same patient. AF555-labelled PEG-MS (2 × 10^7^) and PEG particles (1.5 × 10^8^) with or without BsAb functionalization were subsequently added into the PBMCs suspension in the presence of human plasma and incubated at 37 °C for 1 h. Experiments at lower PEG particle concentration (2 × 10^7^) were also conducted. The dose of PEG particles was set at 1.5 × 10^8^ for optimal visualization of CLL targeting. After incubation, the cells were incubated on ice for 10 mins, washed with FACS buffer (700*g*, 7 min), and phenotyped at 4 °C for 1 h using an antibody cocktail consisting of CD14 AF488 (M5E2, BD), CD20 AF647 (2H7, BioLegend), CD3 BV421 (UCHT1, BioLegend) in titrated concentrations. The cells were subsequently washed with FACS wash buffer (4 mL, 700*g*, 7 min) and PBS (4 mL, 700*g*, 7 min), fixed with 4% paraformaldehyde in PBS (500 μL) for 20 min at 22 °C, then washed with PBS twice (3 mL, 1000*g*, 10 mins), and finally mounted on a 8-well Lab-Tek chambered coverglass slide (Thermo Fisher Scientific, USA). The 8-well Lab-Tek chambered coverglass slide was pre-treated with poly-L-lysine (200 μL, 0.01% w/v, Mw 70−150 kDa) solution for 14 h at 22 °C, followed by washing with PBS and air-drying. The fixed cells were allowed to set in the dark for 12 h at 4 °C prior to imaging. Cells were imaged using a Nikon A1R+ confocal microscope with a 60x/1.4 NA Plan Apo λ oil immersion objective lens and 405nm, 488nm, 561nm, and 640nm lasers.

### Cytotoxicity of nanoparticles in whole blood of CLL patients

The fresh human blood was collected from CLL patients into sodium heparin vacuettes (Greiner Bio-One) and counted with a CELL-DYN Emerald analyzer. The fresh blood (100 μL) was subsequently incubated with DiD-labelled Doxil nanoparticles (2.2 μg based on lipid, equivalent to 0.3 μg doxorubicin) with or without BsAb functionalization at 37 °C for 24 h. Experiments at lower Doxil doses (0.44 μg based on lipid, equivalent to 0.06 μg doxorubicin) or shorter incubation period (3 h) were also conducted to optimize the conditions. The dose of Doxil nanoparticles and the incubation period were set as 2.2 μg based on lipid (equivalent to 0.3 μg doxorubicin) and 24 h for optimal cytotoxicity to CLL cells. After incubation, the whole blood was incubated on ice for 10 mins, and the red blood cells were lysed using Pharm Lyse buffer (4 mL) and removed by centrifugation (500*g*, 5 min). After being washed twice with FACS buffer, the cells were phenotyped at 4 °C for 1 h using an antibody cocktail consisting of CD3 AF700 (SP34-2, BD), CD14 APC-H7 (MΦP9, BD), CD66b BV421 (G10F5, BD), CD56 PE-Cy7 (HCD56, BioLegend), CD19 BV650 (HIB19, BioLegend), CD5 BV786 (UCHT2, BD) and CD22 PE (S-HCL-1, BD) in titrated concentrations. To evaluate the cytotoxicity of Doxil, cells were incubated with SYTOX™ AADvanced™ dead cell stain solution at a final concentration of 1 μM at 22 °C for 5 min, followed by stored on ice and directly analyzed by flow cytometry (LSRFortessa, BD Bioscience). Dead cells were detected using 488 nm excitation and emission collected in a 695/40 bandpass. The data were processed using FlowJo V10. Live cell (%) refers to the proportion of each cell type without fluorescence above background, stemming from SYTOX™ AADvanced™ dead cell stain (See flow cytometry histogram examples in Fig. 6a). Live cell (%) data are shown as the mean of three independent experiments (using the fresh blood from each donor), with at least 400,000 leukocytes analyzed for each experimental condition studied.

### Statistical analysis

Correlation analysis in Fig. 5 and Supplementary Fig. 11−13 were assessed using nonparametric Spearman correlation tests in GraphPad Prism 10.2.2.

### Reporting Summary

Further information on research design is available in the Nature Research Reporting Summary linked to this article.

### Data availability

The main data supporting the results in this study are available within the paper and its Supplementary Information. The raw and analysed datasets generated during the study are too large to be publicly shared, yet they will be made available for research purposes from the corresponding authors on reasonable request.

## Data Availability

All data produced in the present study are available upon reasonable request to the authors

## Acknowledgements

We thank the participations for the generous involvement and provision of samples. We thank David Westerman, David Ritchie, Michael Dickinson, and Ashley Whitechurch (Peter MacCallum cancer centre) and Rachel Koldej (Royal Melbourne Hospital) for help with subject recruitment. We thank Chris B. Howard and Pie Huda (University of Queensland) for providing BsAbs and Thakshila H. Amarasena (University of Melbourne) for excellent technical assistance. We acknowledge the Materials Characterisation and Fabrication Platform (MCFP) at The University of Melbourne for assistance in confocal microscopy and super-resolution structured illumination microscopy. This study was supported by an Australian Research Council (ARC) Discovery Project (DP210103114 to FC, SJK, YJ), an ARC Discovery Early Career Researcher Award (DE230101542 to YJ), a Maxwell Eagle Endowment Award for Cancer Research (to YJ and SJK), an RMIT Vice-Chancellor’s Postdoctoral Fellowship (to YJ), an NHMRC program grant (GNT1149990 to SJK), and NHMRC Investigator grants (SJK, GNT2016732 to FC, KT).

## Author contributions

YJ and SJK. conceived, designed, and supervised the study and drafted the manuscript. YJ, SL, AEQT, EHO, PTB performed experiments and provided technical advice. YJ, AEQT, CT, and SJK recruited subjects and processed their blood samples. MP, JC, FC, and KJT provided intellectual input and reagents. All authors approved the final version of the manuscript.

## References

1) de Lázaro, I.; Mooney, D. J., Obstacles and opportunities in a forward vision for cancer nanomedicine. Nat. Mater. 2021, 20, 1469.

2) Sharma, A.; Jasrotia, S.; Kumar, A., Effects of Chemotherapy on the Immune System: Implications for Cancer Treatment and Patient Outcomes. Naunyn-Schmiedeberg’s Arch. Pharmacol. 2023, DOI: 10.1007/s00210-023-02781-2.

3) Fan, D.; Cao, Y.; Cao, M.; Wang, Y.; Cao, Y.; Gong, T., Nanomedicine in cancer therapy. Signal Transduct. Target. Ther. 2023, 8, 293.

4) Moles, E.; Howard, C. B.; Huda, P.; Karsa, M.; McCalmont, H.; Kimpton, K.; Duly, A.; Chen, Y.; Huang, Y.; Tursky, M. L.; Ma, D.; Bustamante, S.; Pickford, R.; Connerty, P.; Omari, S.; Jolly, C. J.; Joshi, S.; Shen, S.; Pimanda, J. E.; Dolnikov, A.; Cheung, L. C.; Kotecha, R. S.; Norris, M. D.; Haber, M.; de Bock, C. E.; Somers, K.; Lock, R. B.; Thurecht, K. J.; Kavallaris, M., Delivery of PEGylated liposomal doxorubicin by bispecific antibodies improves treatment in models of high-risk childhood leukemia. Sci. Transl. Med. 2023, 15, eabm1262.

5) Mahmoudi, M.; Landry, M. P.; Moore, A.; Coreas, R., The protein corona from nanomedicine to environmental science. Nat. Rev. Mater. 2023, 8, 422.

6) Farshbaf, M.; Valizadeh, H.; Panahi, Y.; Fatahi, Y.; Chen, M.; Zarebkohan, A.; Gao, H., The impact of protein corona on the biological behavior of targeting nanomedicines. Int. J. Pharm. 2022, 614, 121458.

7) Sivaram, A. J.; Wardiana, A.; Alcantara, S.; Sonderegger, S. E.; Fletcher, N. L.; Houston, Z. H.; Howard, C. B.; Mahler, S. M.; Alexander, C.; Kent, S. J.; Bell, C. A.; Thurecht, K. J., Controlling the Biological Fate of Micellar Nanoparticles: Balancing Stealth and Targeting. ACS Nano 2020, 14, 13739.

8) Cui, J.; De Rose, R.; Alt, K.; Alcantara, S.; Paterson, B. M.; Liang, K.; Hu, M.; Richardson, J. J.; Yan, Y.; Jeffery, C. M.; Price, R. I.; Peter, K.; Hagemeyer, C. E.; Donnelly, P. S.; Kent, S. J.; Caruso, F., Engineering Poly(ethylene glycol) Particles for Improved Biodistribution. ACS Nano 2015, 9, 1571.

9) Ju, Y.; Kelly, H. G.; Dagley, L. F.; Reynaldi, A.; Schlub, T. E.; Spall, S. K.; Bell, C. A.; Cui, J.; Mitchell, A. J.; Lin, Z.; Wheatley, A. K.; Thurecht, K. J.; Davenport, M. P.; Webb, A. I.; Caruso, F.; Kent, S. J., Person-Specific Biomolecular Coronas Modulate Nanoparticle Interactions with Immune Cells in Human Blood. ACS Nano 2020, 14, 15723.

10) Ju, Y.; Lee, W. S.; Pilkington, E. H.; Kelly, H. G.; Li, S.; Selva, K. J.; Wragg, K. M.; Subbarao, K.; Nguyen, T. H. O.; Rowntree, L. C.; Allen, L. F.; Bond, K.; Williamson, D. A.; Truong, N. P.; Plebanski, M.; Kedzierska, K.; Mahanty, S.; Chung, A. W.; Caruso, F.; Wheatley, A. K.; Juno, J. A.; Kent, S. J., Anti-PEG Antibodies Boosted in Humans by SARS-CoV-2 Lipid Nanoparticle mRNA Vaccine. ACS Nano 2022, 16, 11769.

11) Cui, J.; Ju, Y.; Houston, Z. H.; Glass, J. J.; Fletcher, N. L.; Alcantara, S.; Dai, Q.; Howard, C. B.; Mahler, S. M.; Wheatley, A. K.; De Rose, R.; Brannon, P. T.; Paterson, B. M.; Donnelly, P. S.; Thurecht, K. J.; Caruso, F.; Kent, S. J., Modulating Targeting of Poly(ethylene glycol) Particles to Tumor Cells Using Bispecific Antibodies. Adv. Healthcare Mater. 2019, 8, 1801607.

12) Barenholz, Y. C., Doxil®—the first FDA-approved nano-drug: lessons learned. J. Controlled Release 2012, 160, 117.

13) Chen, B.-M.; Cheng, T.-L.; Roffler, S. R., Polyethylene Glycol Immunogenicity: Theoretical, Clinical, and Practical Aspects of Anti-Polyethylene Glycol Antibodies. ACS Nano 2021, 15, 14022.

14) Salvati, A.; Pitek, A. S.; Monopoli, M. P.; Prapainop, K.; Bombelli, F. B.; Hristov, D. R.; Kelly, P. M.; Åberg, C.; Mahon, E.; Dawson, K. A., Transferrin-functionalized nanoparticles lose their targeting capabilities when a biomolecule corona adsorbs on the surface. Nat. Nanotechnol. 2013, 8, 137.

15) Yao, X.; Qi, C.; Sun, C.; Huo, F.; Jiang, X., Poly(ethylene glycol) alternatives in biomedical applications. Nano Today 2023, 48, 101738.

16) Nogueira, S. S.; Schlegel, A.; Maxeiner, K.; Weber, B.; Barz, M.; Schroer, M. A.; Blanchet, C. E.; Svergun, D. I.; Ramishetti, S.; Peer, D.; Langguth, P.; Sahin, U.; Haas, H., Polysarcosine-Functionalized Lipid Nanoparticles for Therapeutic mRNA Delivery. ACS Appl. Nano Mater. 2020, 3, 10634.

17) Sanchez, A. J. D. S.; Loughrey, D.; Echeverri, E. S.; Huayamares, S. G.; Radmand, A.; Paunovska, K.; Hatit, M.; Tiegreen, K. E.; Santangelo, P. J.; Dahlman, J. E., Substituting Poly(ethylene glycol) Lipids with Poly(2-ethyl-2-oxazoline) Lipids Improves Lipid Nanoparticle Repeat Dosing. Adv. Healthcare Mater. 2024, DOI: 10.1002/adhm.2023040332304033.

18) Rajesh, S.; Leiske, M. N.; Leitch, V.; Zhai, J.; Drummond, C. J.; Kempe, K.; Tran, N., Lipidic poly(2-oxazoline)s as PEG replacement steric stabilisers for cubosomes. J. Colloid Interface Sci. 2022, 623, 1142.

19) Qiao, R.; Fu, C.; Li, Y.; Qi, X.; Ni, D.; Nandakumar, A.; Siddiqui, G.; Wang, H.; Zhang, Z.; Wu, T.; Zhong, J.; Tang, S.-Y.; Pan, S.; Zhang, C.; Whittaker, M. R.; Engle, J. W.; Creek, D. J.; Caruso, F.; Ke, P. C.; Cai, W.; Whittaker, A. K.; Davis, T. P., Sulfoxide-Containing Polymer-Coated Nanoparticles Demonstrate Minimal Protein Fouling and Improved Blood Circulation. Adv. Sci. 2020, 7, 2000406.

20) Ibrahim, M.; Shimizu, T.; Ando, H.; Ishima, Y.; Elgarhy, O. H.; Sarhan, H. A.; Hussein, A. K.; Ishida, T., Investigation of anti-PEG antibody response to PEG-containing cosmetic products in mice. J. Controlled Release 2023, 354, 260.

21) Gaballa, S. A.; Shimizu, T.; Takata, H.; Ando, H.; Ibrahim, M.; Emam, S. E.; Amorim Matsuo, N. C.; Kim, Y.; Naguib, Y. W.; Mady, F. M.; Khaled, K. A.; Ishida, T., Impact of Anti-PEG IgM Induced via the Topical Application of a Cosmetic Product Containing PEG Derivatives on the Antitumor Effects of PEGylated Liposomal Antitumor Drug Formulations in Mice. Mol. Pharm. 2024, 21, 622.

22) Ju, Y.; Carreño, J. M.; Simon, V.; Dawson, K.; Krammer, F.; Kent, S. J., Impact of anti-PEG antibodies induced by SARS-CoV-2 mRNA vaccines. Nat. Rev. Immunol. 2023, 23, 135.

23) Li, S.; Ma, Y.; Cui, J.; Caruso, F.; Ju, Y., Engineering poly(ethylene glycol) particles for targeted drug delivery. Chem. Commun. 2024, 60, 2591.

24) Ju, Y.; Kim, C.-J.; Caruso, F., Functional Ligand-Enabled Particle Assembly for Bio–Nano Interactions. Acc. Chem. Res. 2023, 56, 1826−1837.

25) Duggan, S. T.; Keating, G. M., Pegylated Liposomal Doxorubicin. Drugs 2011, 71, 2531.

