## Supporting Information for "Ex vivo human leukemia blood model illustrates limitations of cancer-targeting PEGylated nanoparticles"

### Methods

**Preparation and characterization of BsAb-conjugated particles.** Alex Fluor 647 (AF647) or AF555-labelled PEGylated mesoporous silica (PEG-MS) and PEG particles were prepared using a previously published method.[1] Dioctadecyl-3,3,3,3-tetramethylindodicarbocyanine (DiD)-labeled PEGylated doxorubicin-encapsulated liposomes that were formulated with the same lipid composition and drug/lipid ratio as FDA-approved liposomal doxorubicin agent (Doxil®), in addition to 0.2 mol % DiD, were purchased from FormuMax Scientific Inc., USA and named as Doxil nanoparticles. The BsAbs were synthesized based on the previously published method.[2] The functionalization of BsAb on particles was based on a previously published method with slight modification.[3] Briefly, PEG particles ($5\times10^9$), PEG-MS particles ($10^9$), or Doxil (100 ug based on lipid concentration) were incubated with 50 μg BsAb in PBS at 4 ℃ under orbital mixing for 15 h. Transmission electron microscopy (TEM) images of PEG and PEG-MS particles were acquired using an FEI Tecnai TF20 instrument. PEG and PEG-MS particle suspensions were dropped and air-dried on Formvar carbon-coated copper grids (plasma-treated). Cryogenic TEM (Cryo-TEM) images of Doxil particles were acquired using a FEI Tecnai Spirit instrument at an operation voltage of 120 kV under liquid nitrogen cooling under low dose conditions. Doxil suspensions were applied to glow-discharged lacey carbon grids and vitrified using a Vitrobot (FEI) system. Vitrified sample grids were stored in liquid nitrogen prior to imaging. Super-resolution structured illumination microscopy (SIM) of the particles was performed on a Zeiss Elyra 7 Lattice SIM with a 63x/1.4 NA Plan-Apochromat oil immersion objective lens, a pco.edge sCMOS 4.2 CL HS camera, and a 642nm excitation laser. Image acquisition and SIM reconstruction were performed with Zeiss ZEN Black 3.1 SR software. Dynamic light scattering (DLS) analysis of the particles was performed on a Zetasizer Nano-ZS (Malvern Instruments, UK) instrument. Zeta-potential measurements of the particles were performed at pH 7.4 in phosphate buffer (5 mM) using a Zetasizer Nano-ZS (Malvern Instruments, UK). Particle counting for PEG and PEG-MS particles was performed using an Apogee A50-Micro flow cytometer (Apogee Flow Systems, UK). The concentration of Doxil (based on lipid) was provided by the manufacturer.

**Evaluation of cancer cell targeting using cell lines in vitro.** Raji, Jurkat, Hut-78 and THP-1 cell lines were purchased from ATCC (USA) and maintained in complete RPMI-1640 media containing 10% FCS and GlutaMAX at 37 ℃ in a cell culture incubator with 5 % CO2 and 95% relative humidity. To assess cell targeting, Raji, Jurkat, Hut-78 and THP-1 cells ($1 \times 10^5$) were incubated with fluorescence-labelled PEG ($2 \times 10^7$), PEG-MS ($2 \times 10^7$) and Doxil nanoparticles (0.44 μg based on lipid) with or without BsAb functionalization in complete RPMI-1640 media (100 μL) at 37 ℃ for 1 h. To assess the CD20 and CD28 expression, CD20 AF647 (2H7, BioLegend) or CD28 AF647 (CD28.2, BioLegend) were incubated with cells at 4 °C for 1 h. After incubation, the cells were incubated on ice for 10 mins, washed with flow cytometry staining (FACS) wash buffer (PBS containing 2 mM EDTA and 0.5% w/v BSA) twice, and analyzed by flow cytometry (LSRFortessa, BD Bioscience). The data were processed using FlowJo V10. The degree of cell association of the particles was evaluated by using the percentage of cells that exhibited stronger fluorescence intensity than control cells (untreated cells). Data were reported as a mean of three independent experiments with at least 40,000 cells analyzed for each experimental condition studied.

**Anti-PEG Antibody ELISA.** The ELISA to detect anti-PEG IgG and IgM was conducted used a previously developed method.[4] Briefly, the 8-arm PEG-$NH_2$ (40 kDa, 200 μg $mL^{-1}$, JenKem Technology, USA) in PBS was coated onto MaxiSorp 96-well plates (Nunc, Denmark) for 18 h at 4 °C, followed by washing with PBS four times. Plates were blocked with 5% (w/v) skim milk powder in PBS at 4 °C for 22 h, followed by adding serially diluted human plasma in 5% skim milk in duplicate for 1 h at 22 °C. Plates were washed with 0.1% 3-[(3-cholamidopropyl)-dimethylammonio]-1-propanesulfonate (CHAPS, Sigma-Aldrich, USA)/PBS buffer twice and PBS four times prior to addition of an HRP-conjugated anti-human IgG (Dako Agilent, USA) at 1:20,000 dilution or HRP-conjugated anti-human IgM (Jackson ImmunoResearch Laboratories, USA) at 1:10,000 for 1 h at 22 °C. Plates were washed as above and then developed using 3,3',5,5'-Tetramethylbenzidine (TMB) liquid substrate (Sigma-Aldrich, USA).

Reaction was stopped with 0.16M $H_2SO_4$ and read at 450nm. Endpoint titres were calculated as the reciprocal plasma dilution giving signal 2× background using a fitted curve (4-parameter log regression) and reported as a mean of duplicates. Background was detected by adding the diluted plasma samples (at 1:10 dilution in 5% skim milk) to the non-PEG-coated wells, followed by the same ELISA procedure.

### Supplementary figures and tables

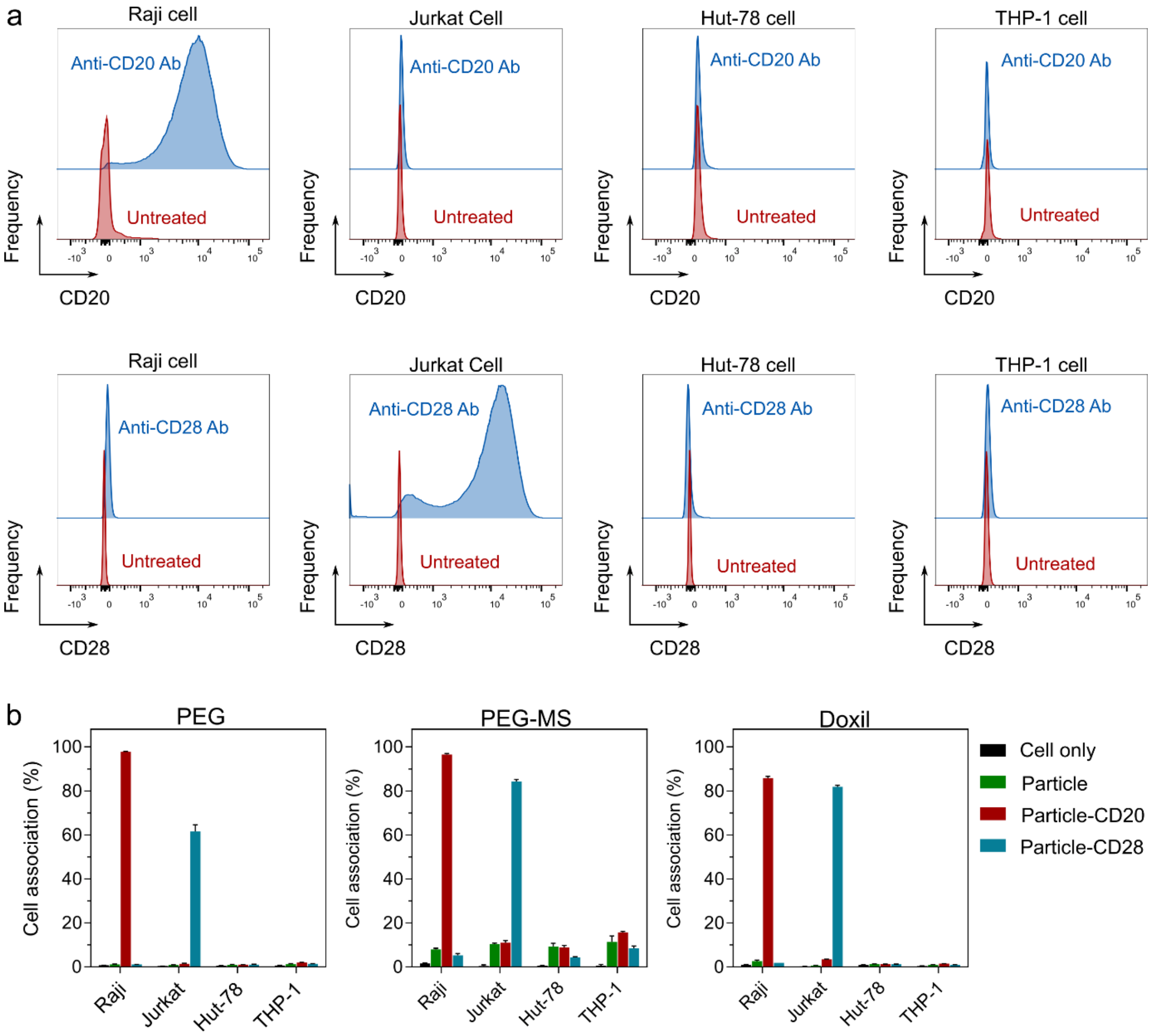


**Supplementary Fig. 1.** Evaluation of cancer cell targeting using conventional cell line models. a) CD20 and CD28 expression of Raji, Jurkat, Hut-78, and THP-1 cell lines. Cells were treated with fluorescence-labelled anti-human CD20 or CD28 antibodies (Ab), followed by analysis by flow cytometry. b) Assessing cell targeting of BsAb-functionalized PEG, PEG-MS, and Doxil nanoparticles using four cell lines: Raji, Jurkat, Hut-78, and THP-1 cells. Particles were incubated with cells for 1 h at 37 ℃, followed by analysis by flow cytometry. Cell association (%) refers to the proportion of each cell type with positive fluorescence, above background, stemming from fluorescence-labeled particles. Cell association (%) data are shown as the mean of three independent experiments (using the fresh blood from the same donor), with at least 40,000 cells analyzed for each experimental condition studied. Cell only control groups represent the respective cell populations without particle incubation.

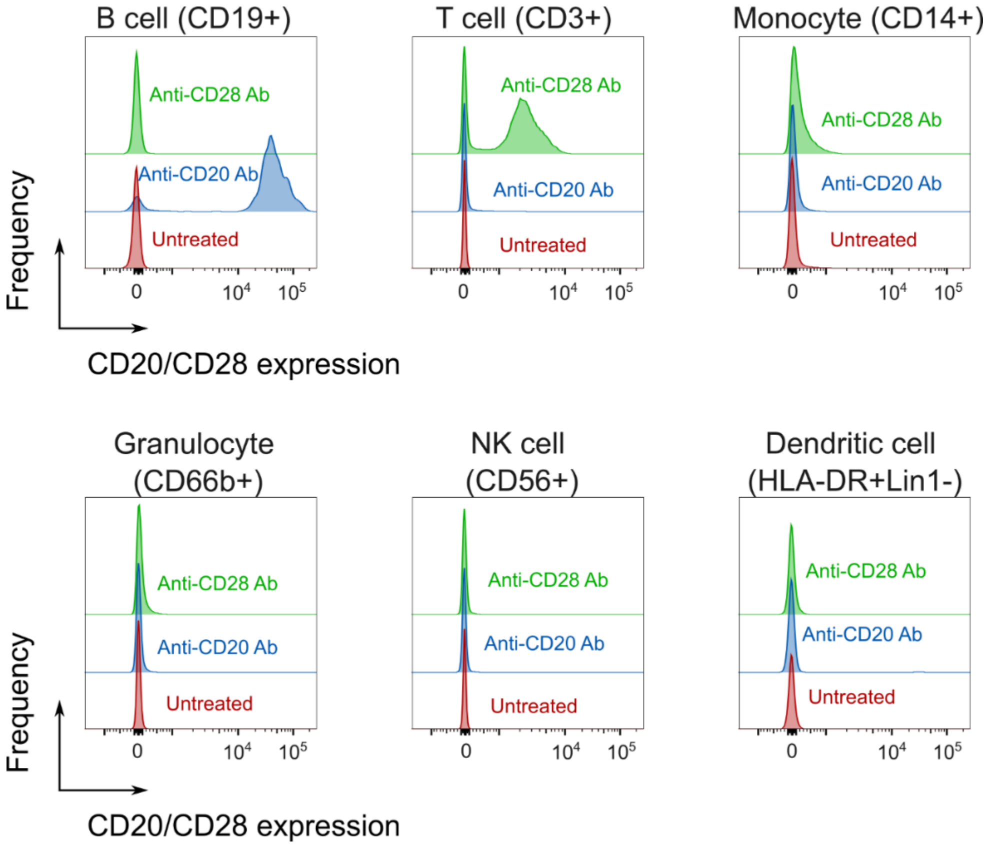


**Supplementary Fig. 2.** Expression of CD20 and CD28 on blood immune cells from a healthy donor. Fresh human blood was treated with fluorescence-labelled anti-human CD20 or CD28 Ab, followed by phenotyping cells with antibody cocktails and analysis by flow cytometry.

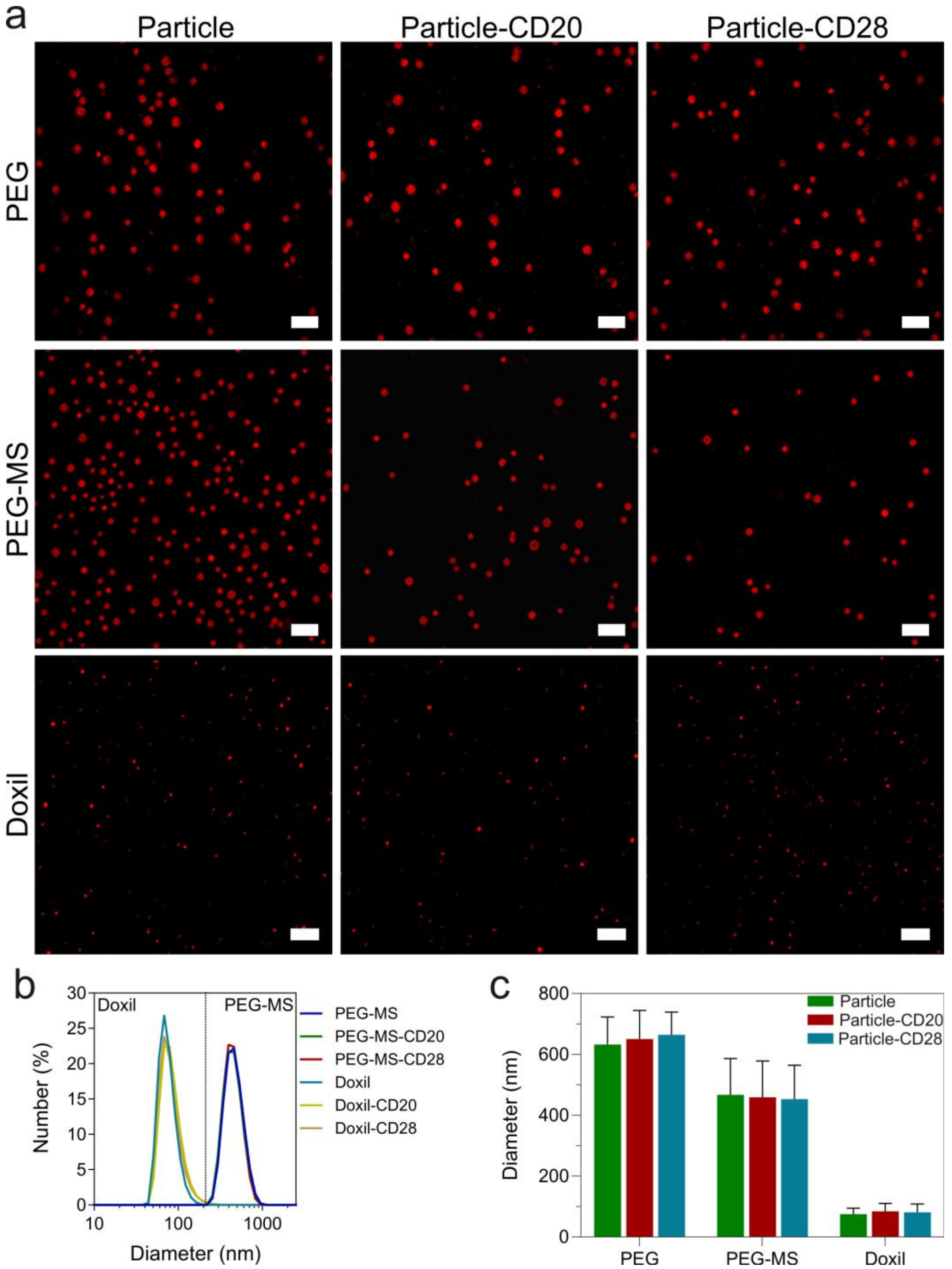


**Supplementary Fig. 3.** Characterization of PEG, PEG-MS, Doxil nanoparticles. (a) Super-resolution structured illumination microscopy (SIM) images of the three types of particles with and without functionalization of anti-PEG/anti-CD20 or anti-PEG/anti-CD28 bispecific antibodies (BsAbs). Scale bars are 2 μm (PEG and PEG-MS) and 1 μm (Doxil), respectively. (b) Dynamic light scattering (DLS) of PEG-MS and Doxil nanoparticles and (c) diameter of the three particles before and after functionalized of BsAbs. Sizes of PEG-MS and Doxil nanoparticles were determined by DLS, presented as the mean ± standard deviation (SD) of three independent measurements. As PEG particles have negligible light scattering, the sizes of the particles were determined by SIM images, shown as the mean ± SD (n = 30). No significant difference in size of each particle before and after functionalization of BsAbs (two-way ANOVA with Tukey's multiple comparisons test) was observed.

**Supplementary Table 1.** Average size and zeta potential of PEG, PEG-MS, and Doxil particles before and after functionalized of BsAbs.

| Particle | Size (nm)[a] | Polydispersity Index[a] | Zeta Potential (mV)[b] |
|---|---|---|---|
| PEG | 632 ± 91 | - | −1 ± 6 |
| PEG-CD20 | 650 ± 94 | - | −1 ± 3 |
| PEG-CD28 | 664 ± 75 | - | -1 ± 4 |
| PEG-MS | 467 ± 119 | 0.017 | -31 ± 5 |
| PEG-MS-CD20 | 459 ± 119 | 0.010 | -27 ± 5 |
| PEG-MS-CD28 | 453 ± 111 | 0.031 | -29 ± 4 |
| Doxil | 75 ± 19 | 0.048 | -13 ± 9 |
| Doxil-CD20 | 84 ± 26 | 0.085 | -9 ± 6 |
| Doxil-CD28 | 81 ± 27 | 0.13 | -10 ± 6 |

[a]The size and polydispersity index of PEG-MS and Doxil particles were determined by dynamic light scattering. The size of PEG particles was determined from the structured illumination microscopy (SIM) images.

[b]Zeta potential measurements were preformed at pH 7.2 in phosphate buffer (5 mM).

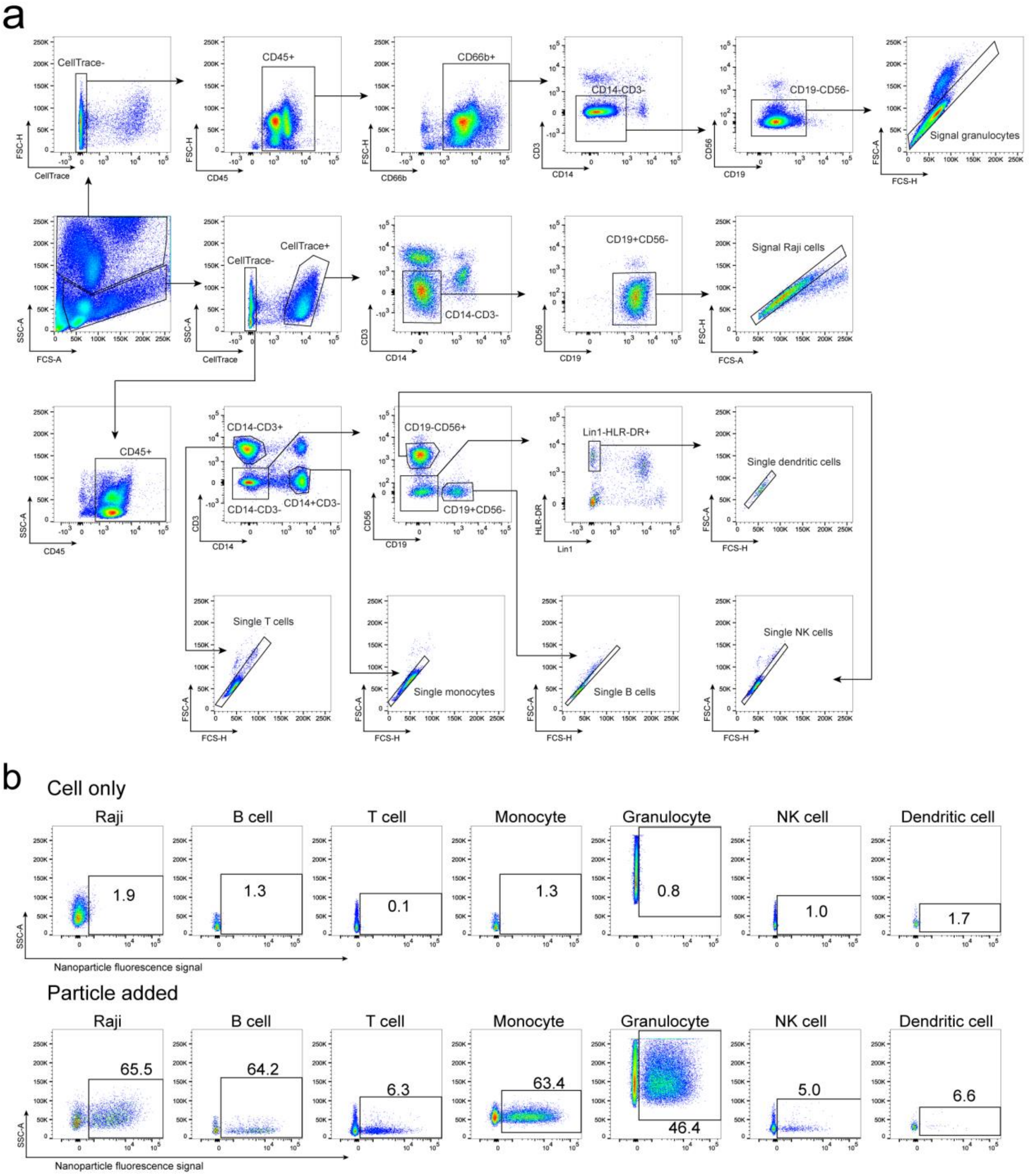


**Supplementary Fig. 4**. Gating strategy used to identify Raji cells and white blood cell population of the whole human blood model that spikes Raji cells into fresh whole blood of a healthy donor. a) Flow cytometry gating strategy used to identify Raji cells and different immune cell populations from human blood. Raji cells were pre-labeled with CellTrace™ Yellow Cell Proliferation Kit and then added into fresh human blood from a healthy donor, followed by incubating with nanoparticles for 1 h at 37 ℃ and subsequent analysis by flow cytometry. b) The percentage of each cell type positive for the fluorescence-labelled nanoparticles was then measured as the cell association (%). The representative gating strategy and cell association percentages for each cell type is shown.

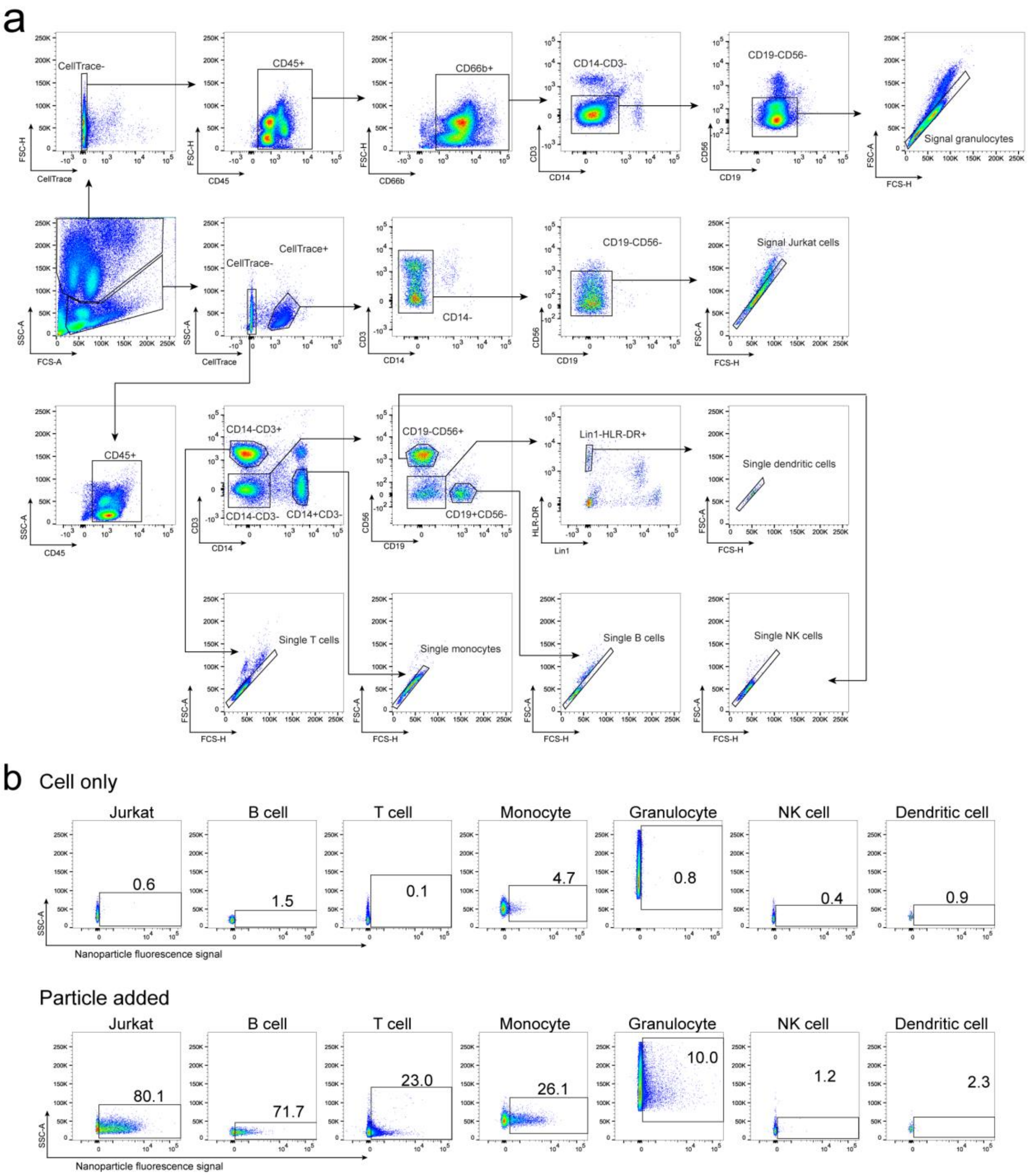


**Supplementary Fig. 5**. Gating strategy used to identify Jurkat cells and white blood cell population of the whole human blood model that spikes Jurkat cells into fresh whole blood of a healthy donor. a) Flow cytometry gating strategy used to identify Jurkat cells and different immune cell populations from human blood. Jurkat cells were pre-labeled with CellTrace™ Yellow Cell Proliferation Kit and then added into fresh human blood from a healthy donor, followed by incubating with nanoparticles for 1 h at 37 ℃ and subsequent analysis by flow cytometry. b) The percentage of each cell type positive for the fluorescence-labelled nanoparticles was then measured as the cell association (%). The representative gating strategy and cell association percentages for each cell type is shown.

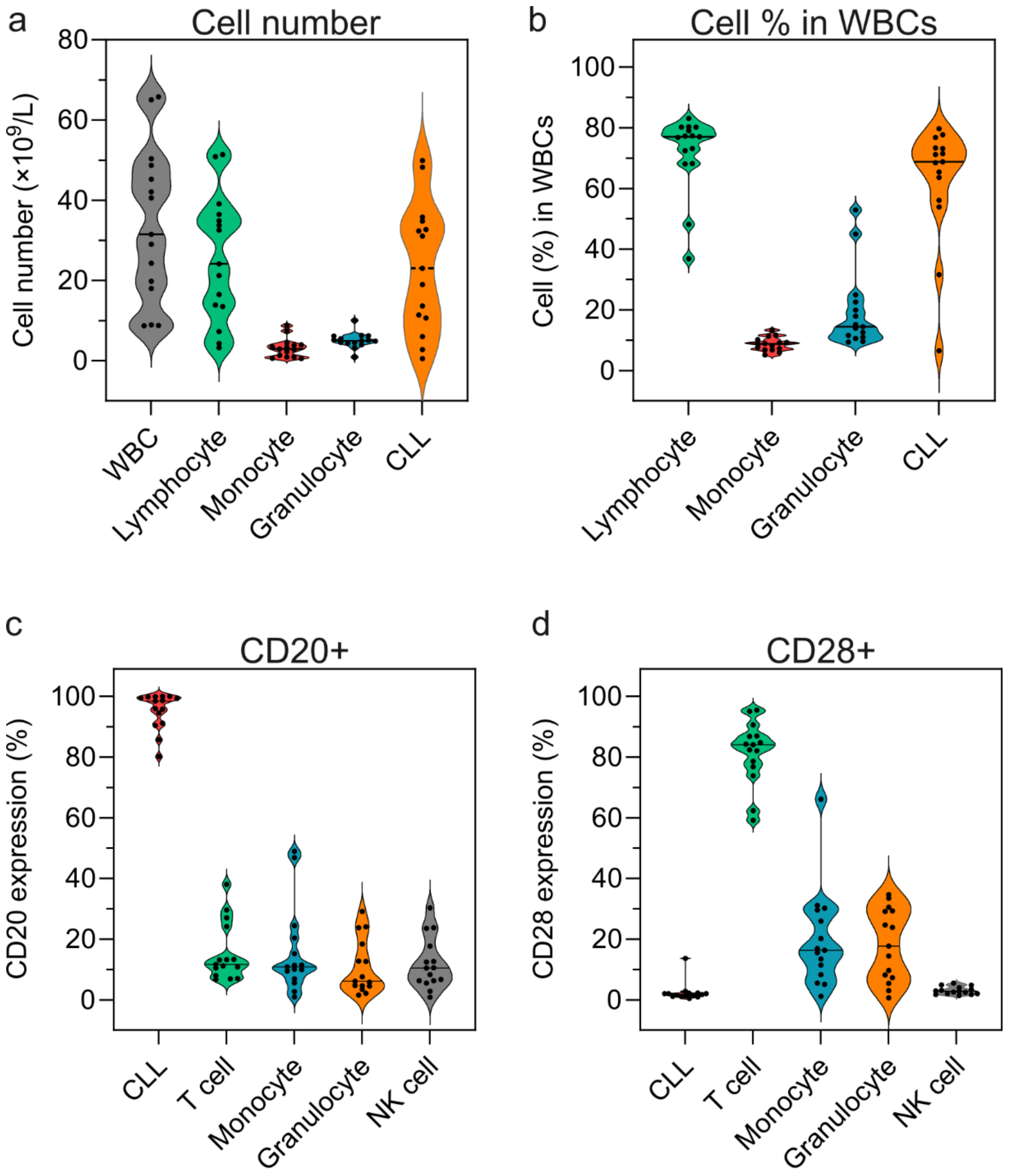


**Supplementary Fig. 6.** Cell count and the expression of CD20 and CD28 on blood immune cells from CLL patients (n=15). a,b) Cell count and cell percentage in white blood cells were obtained from a CELL-DYN Emerald analyzer. CLL cell count was obtained from flow cytometry. c,d) CD20 and CD28 expression of white blood cells from CLL patients. Fresh CLL blood was treated with fluorescence-labelled anti-human CD20 or CD28 Ab, followed by phenotyping cells with antibody cocktails and analysis by flow cytometry.

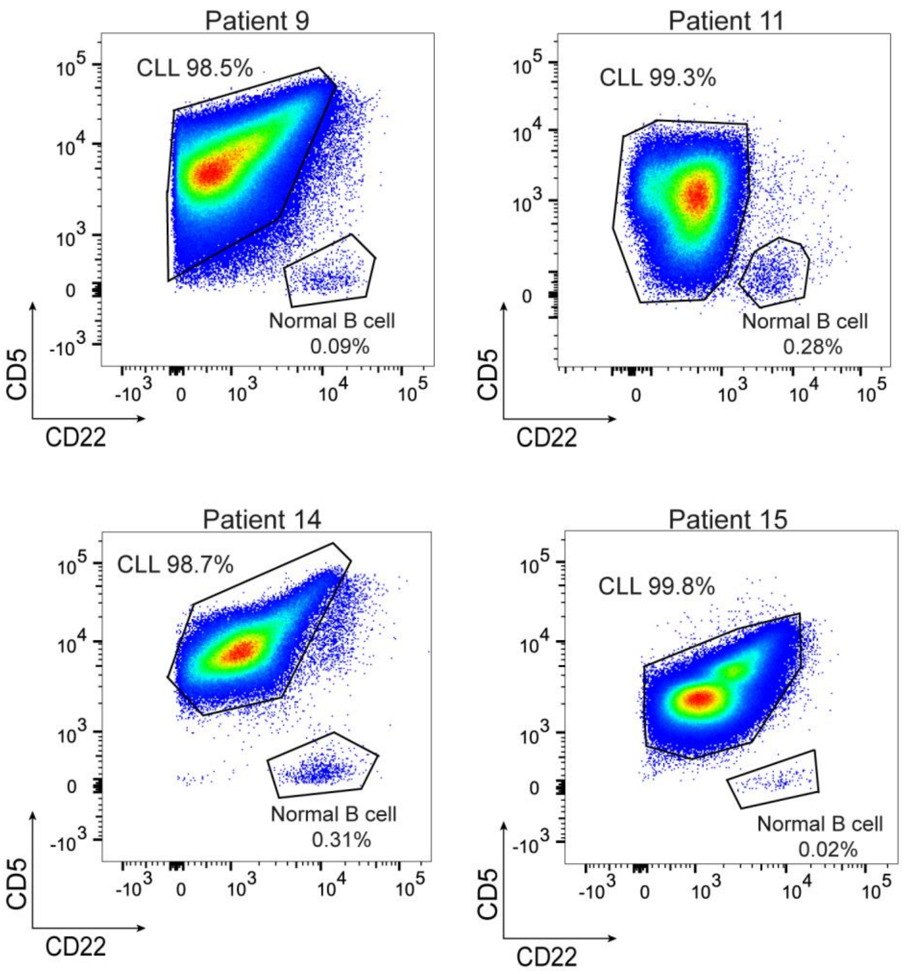


**Supplementary Fig. 7**. Gating strategy used to distinguish between CLL and normal B cells. Only 4 CLL patients have normal B cells identified in the blood with less than 0.5% of total CD19+ B cells. The gating strategy was established based on the previous report on the immunophenotypic profiles of CLL and normal B cells.[5]

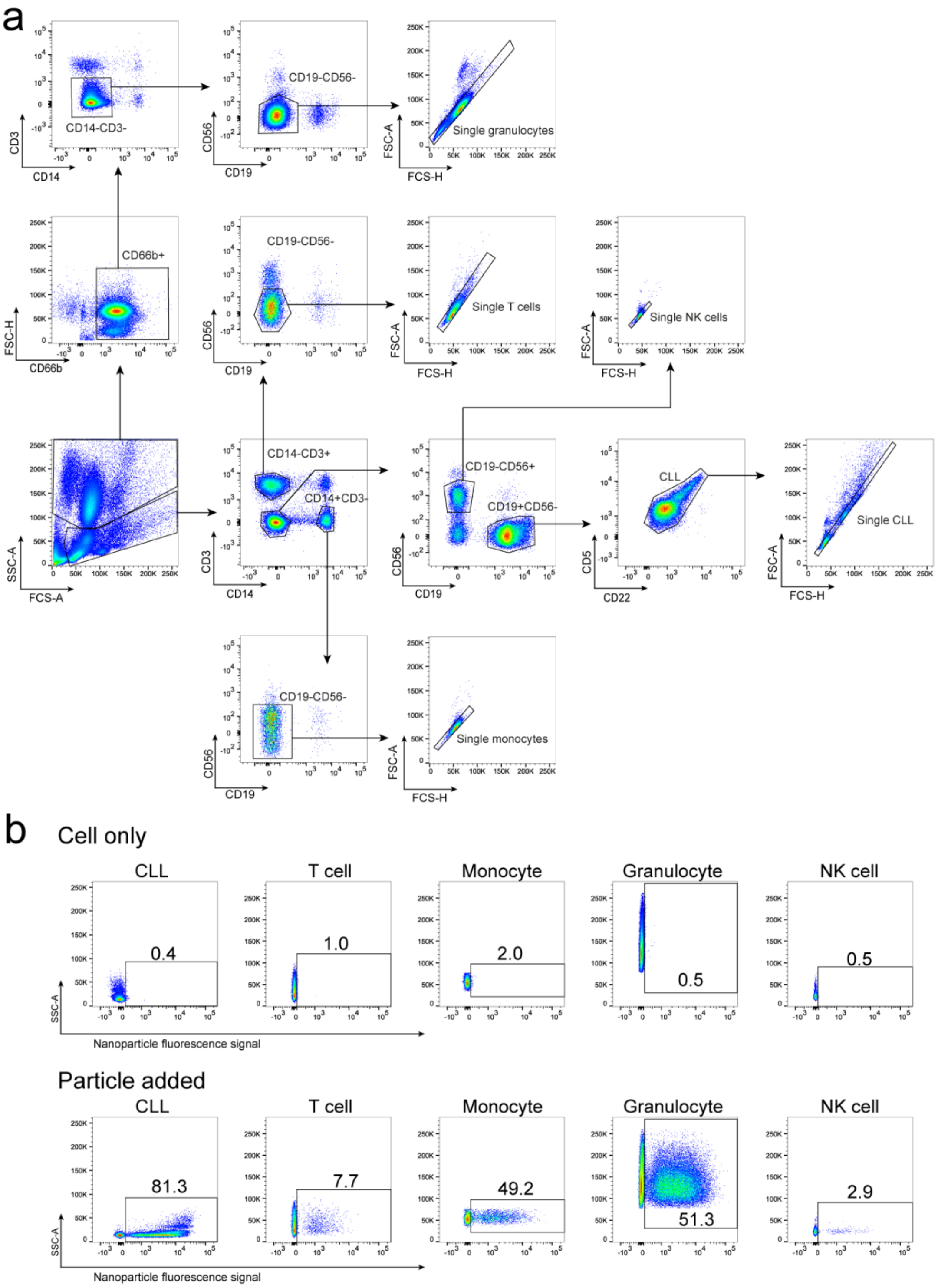


**Supplementary Fig. 8**. Gating strategy used to identify CLL cells and white blood cell population of the whole blood from CLL patients. a) Flow cytometry gating strategy used to identify CLL cells and different immune cell populations from blood of CLL patients. b) The percentage of each cell type positive for the fluorescence-labelled nanoparticles was then measured as the cell association (%). The representative gating strategy and cell association percentages for each cell type is shown.

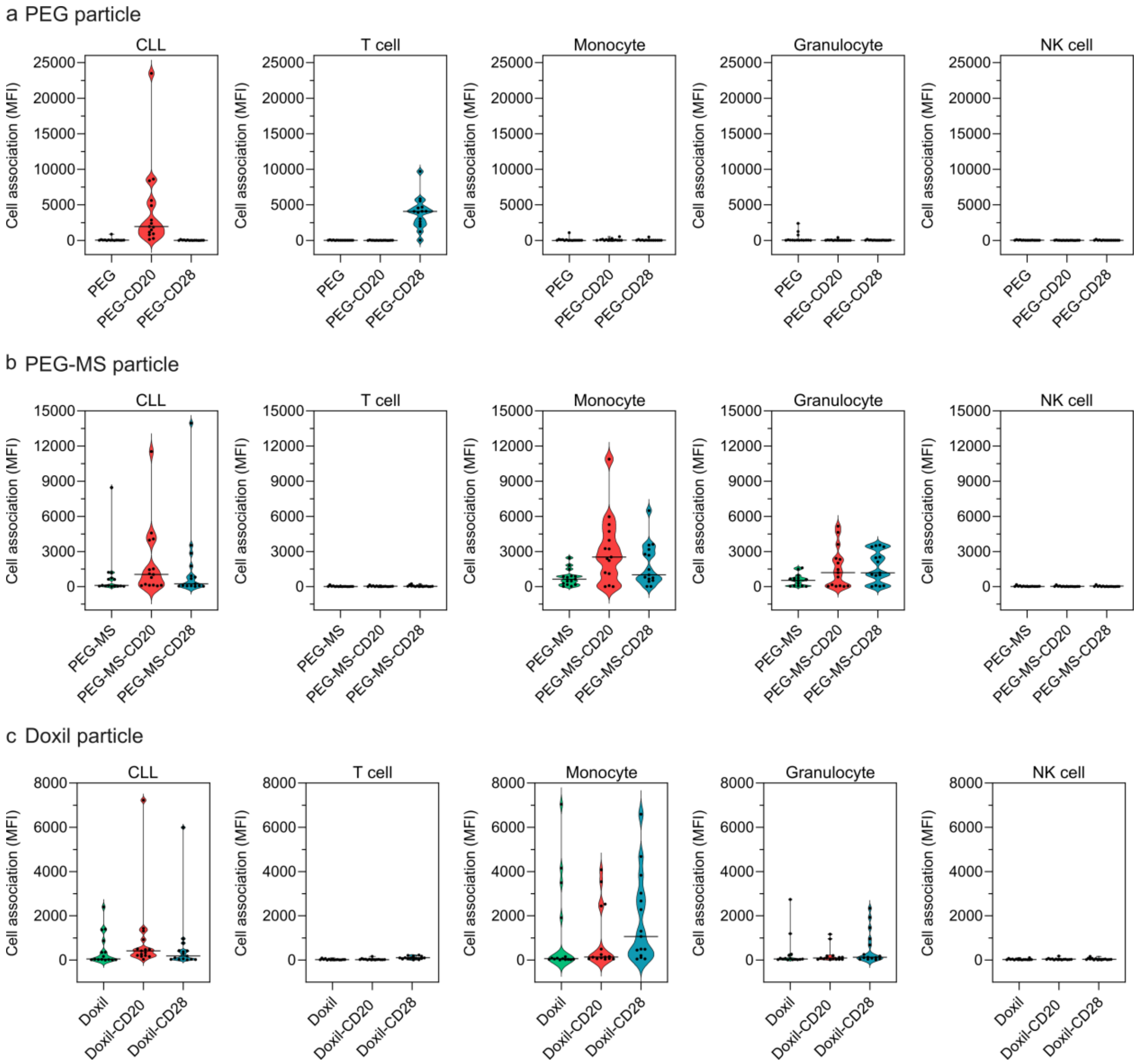


**Supplementary Fig. 9.** Violin plots summarizing cell association (MFI) of the BsAb-functionalized PEG, PEG-MS, and Doxil nanoparticle in 15 CLL patients' blood after 1 h incubation at 37 ℃. Cell association (MFI) refers to the median fluorescence index of each cell type, stemming from fluorescence-labeled particles. Each data point is the mean of three independent experiments (using the same batch of fresh blood from each donor). The median cell association (MFI) across 15 CLL patients are shown as solid lines in the violin plots.

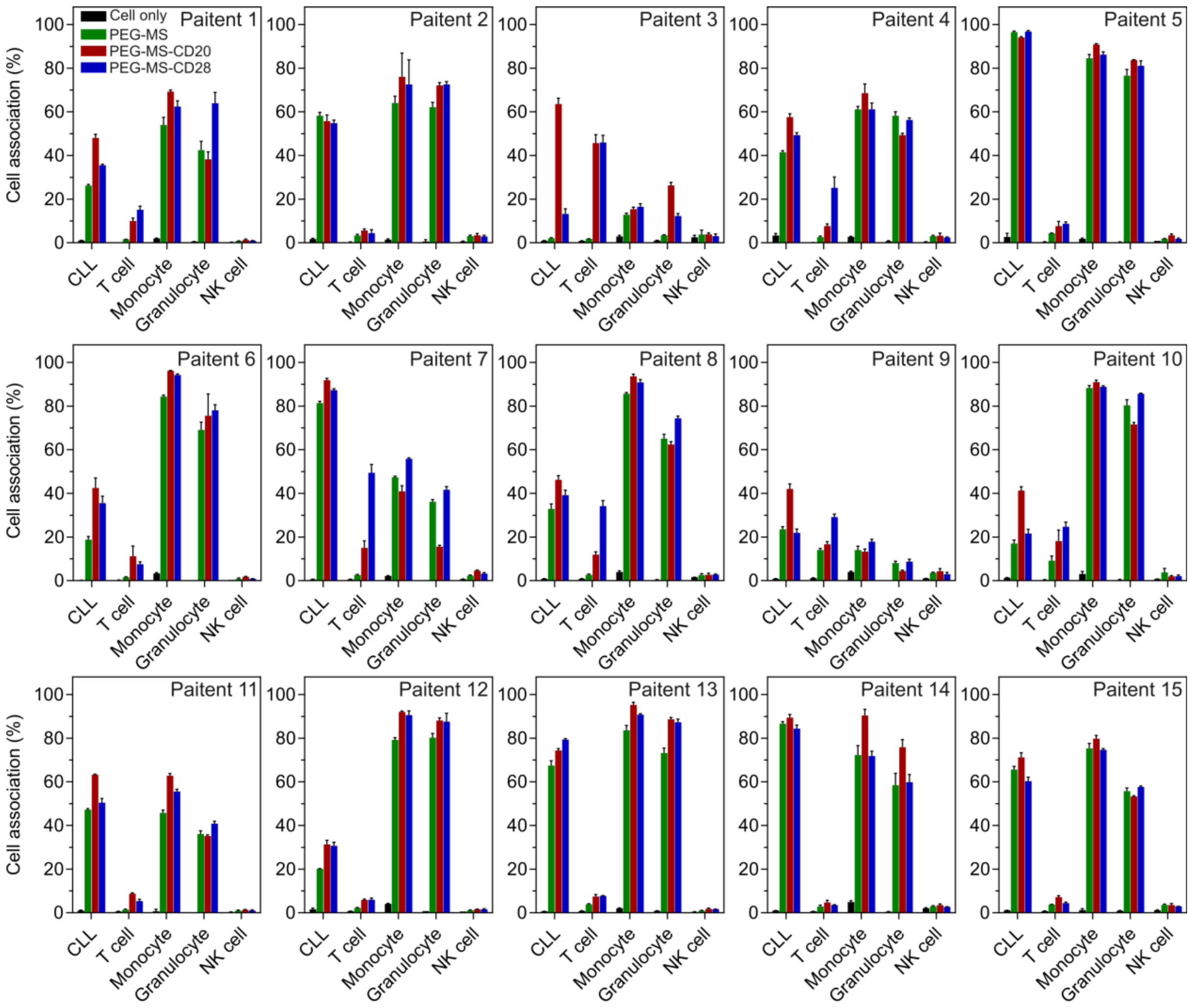


**Supplementary Fig. 10.** CLL targeting of BsAb-functionalized PEG-MS particles in the whole blood of the 15 CLL patients. Cell association (%) refers to the proportion of each cell type with positive fluorescence, above background, stemming from fluorescence-labeled particles (See gating strategy in Supplementary Fig. 8). Cell association (%) data are shown as the mean of three independent experiments (using the fresh blood from each donor), with at least 400,000 leukocytes analyzed for each experimental condition studied. Cell only control groups represent the respective cell populations without particle incubation.

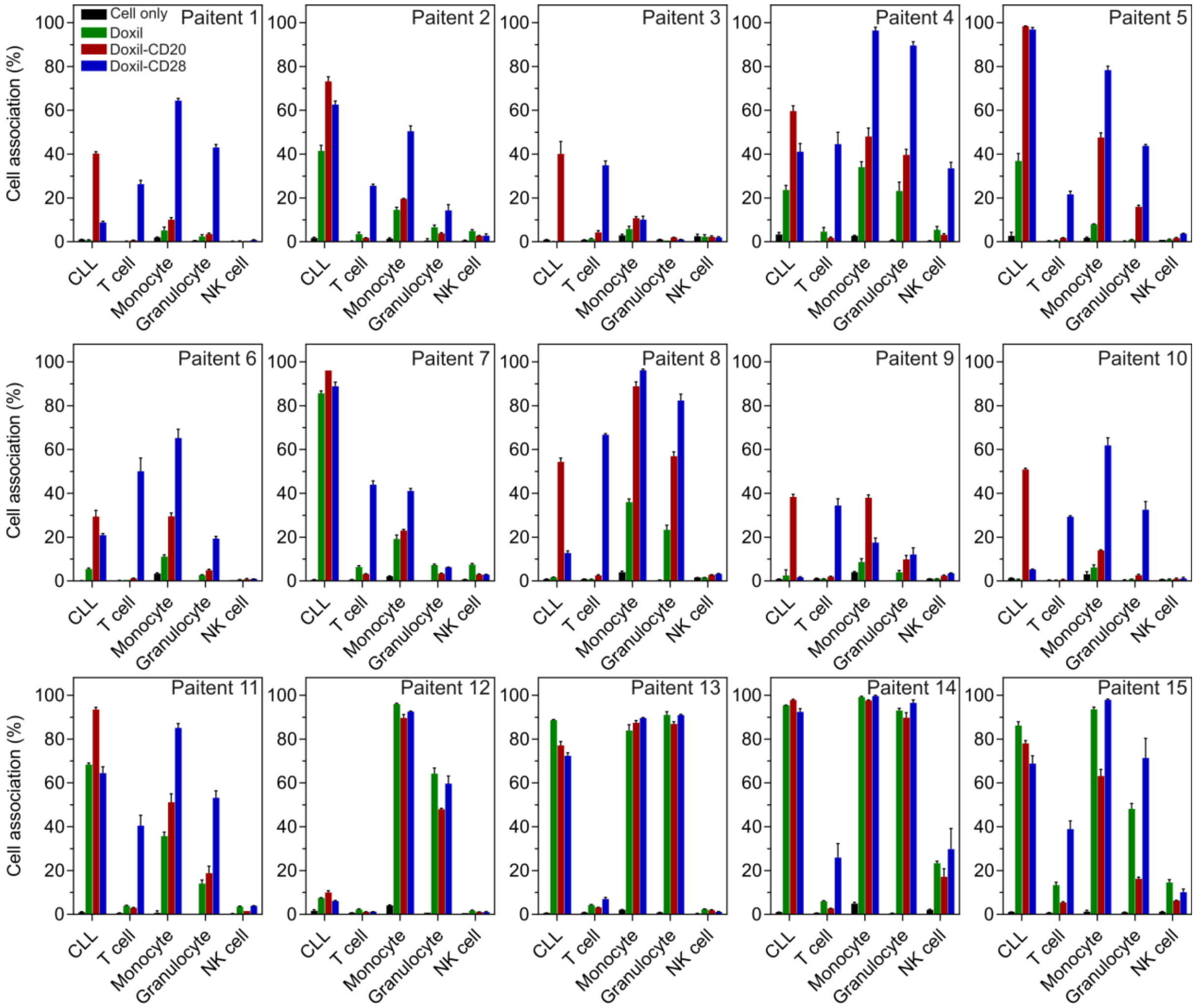


**Supplementary Fig. 11.** CLL targeting of BsAb-functionalized Doxil nanoparticles in the whole blood of the 15 CLL patients. Cell association (%) refers to the proportion of each cell type with positive fluorescence, above background, stemming from fluorescence-labeled particles (See gating strategy in Supplementary Fig. 8). Cell association (%) data are shown as the mean of three independent experiments (using the fresh blood from each donor), with at least 400,000 leukocytes analyzed for each experimental condition studied. Cell only control groups represent the respective cell populations without particle incubation.

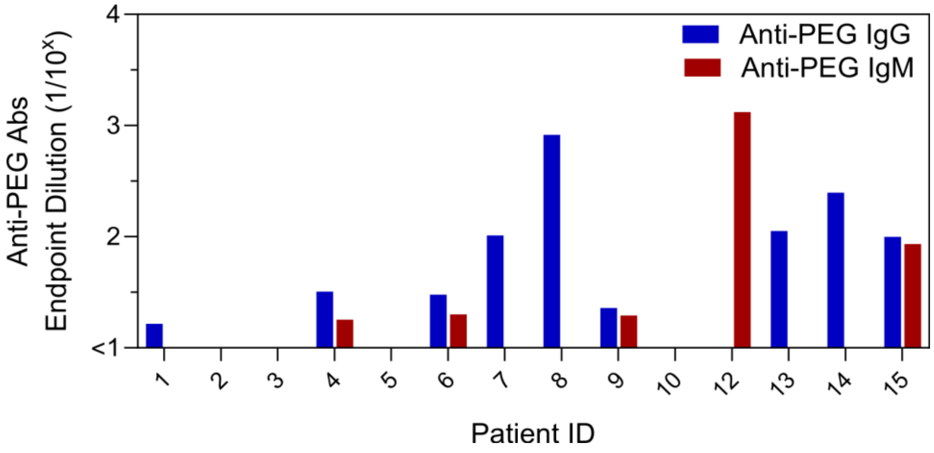


**Supplementary Fig. 12.** Plasma anti-PEG IgG and IgM titers of 14 CLL patients. Data are shown as mean of two independent measurements (Patient 11 was recruited later and thus not included in the assay).

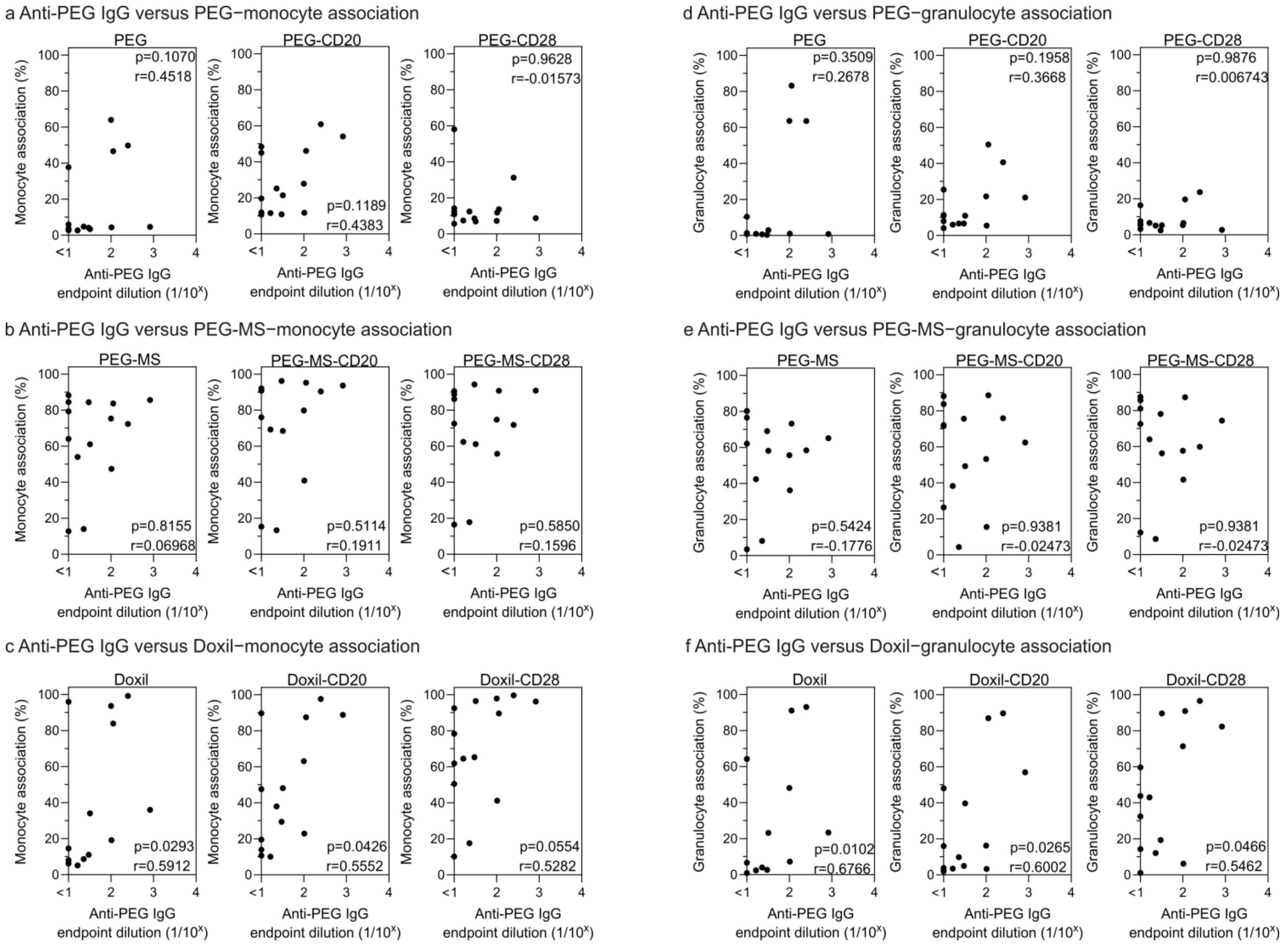


**Supplementary Fig. 13.** Spearman correlation analysis between plasma anti-PEG IgG titters and (a-c) monocyte or (d-f) granulocyte association (%) with PEG, PEG-MS, and Doxil particles with and without functionalization of anti-PEG/anti-CD20 or anti-PEG/anti-CD28 BsAb (n=14).

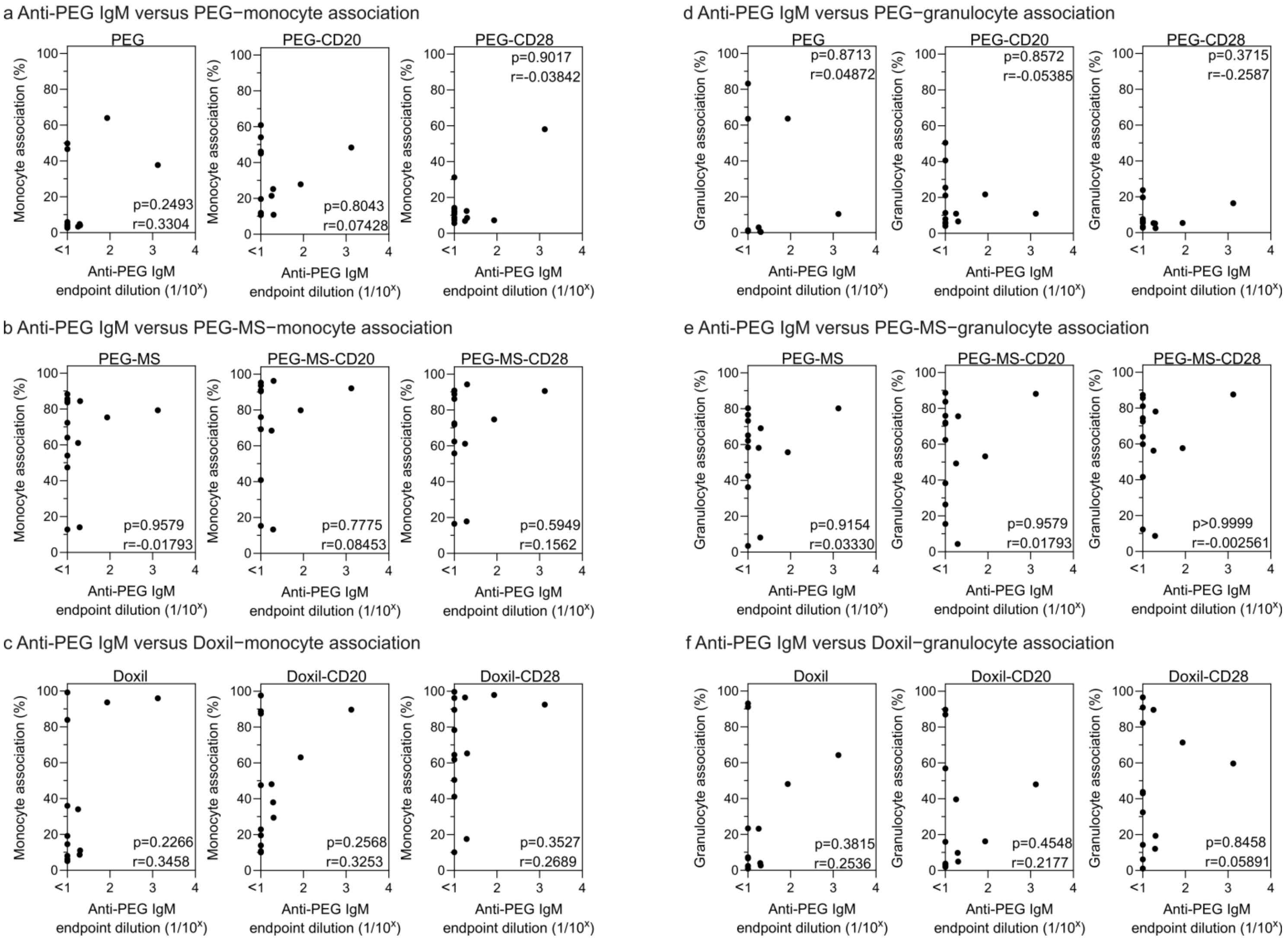


**Supplementary Fig. 14.** Spearman correlation analysis between plasma anti-PEG IgM titters and (a-c) monocyte or (d-f) granulocyte association (%) with PEG, PEG-MS, and Doxil particles with and without functionalization of anti-PEG/anti-CD20 or anti-PEG/anti-CD28 BsAb (n=14).

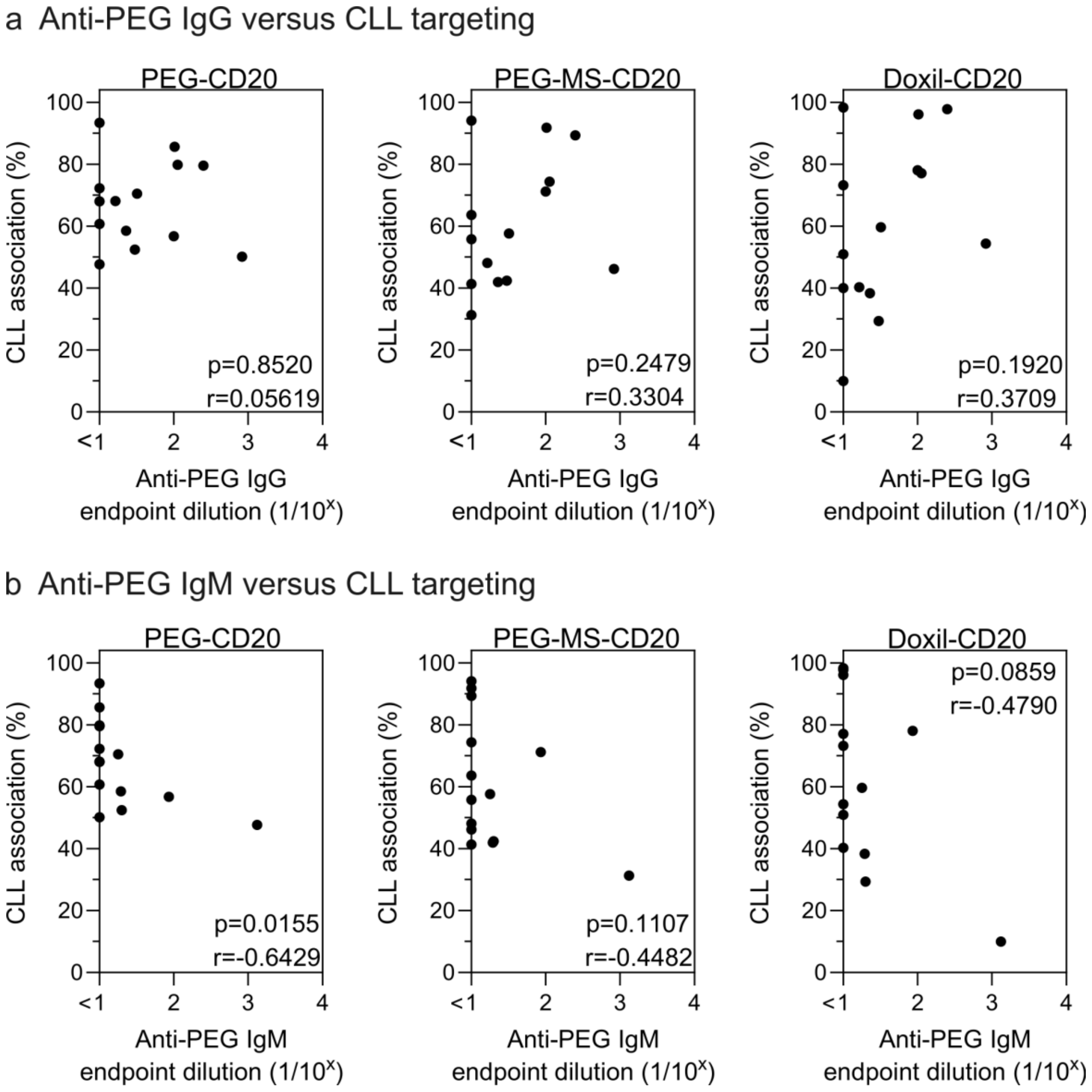


**Supplementary Fig. 15.** Spearman correlation analysis between plasma anti-PEG IgG or IgM titters and CLL targeting of PEG, PEG-MS, and Doxil particles functionalized with anti-PEG/anti-CD20 or anti-PEG/anti-CD28 BsAb (n=14).

**Reference**


1) Ju, Y.; Kelly, H. G.; Dagley, L. F.; Reynaldi, A.; Schlub, T. E.; Spall, S. K.; Bell, C. A.; Cui, J.; Mitchell, A. J.; Lin, Z.; Wheatley, A. K.; Thurecht, K. J.; Davenport, M. P.; Webb, A. I.; Caruso, F.; Kent, S. J., Person-Specific Biomolecular Coronas Modulate Nanoparticle Interactions with Immune Cells in Human Blood. *ACS Nano* **2020,** *14,* 15723.

2) Moles, E.; Howard, C. B.; Huda, P.; Karsa, M.; McCalmont, H.; Kimpton, K.; Duly, A.; Chen, Y.; Huang, Y.; Tursky, M. L.; Ma, D.; Bustamante, S.; Pickford, R.; Connerty, P.; Omari, S.; Jolly, C. J.; Joshi, S.; Shen, S.; Pimanda, J. E.; Dolnikov, A.; Cheung, L. C.; Kotecha, R. S.; Norris, M. D.; Haber, M.; de Bock, C. E.; Somers, K.; Lock, R. B.; Thurecht, K. J.; Kavallaris, M., Delivery of PEGylated liposomal doxorubicin by bispecific antibodies improves treatment in models of high-risk childhood leukemia. *Sci. Transl. Med.* **2023,** *15,* eabm1262.

3) Cui, J.; Ju, Y.; Houston, Z. H.; Glass, J. J.; Fletcher, N. L.; Alcantara, S.; Dai, Q.; Howard, C. B.; Mahler, S. M.; Wheatley, A. K.; De Rose, R.; Brannon, P. T.; Paterson, B. M.; Donnelly, P. S.; Thurecht, K. J.; Caruso, F.; Kent, S. J., Modulating Targeting of Poly(ethylene glycol) Particles to Tumor Cells Using Bispecific Antibodies. *Adv. Healthcare Mater.* **2019,** *8,* 1801607.

4) Ju, Y.; Lee, W. S.; Pilkington, E. H.; Kelly, H. G.; Li, S.; Selva, K. J.; Wragg, K. M.; Subbarao, K.; Nguyen, T. H. O.; Rowntree, L. C.; Allen, L. F.; Bond, K.; Williamson, D. A.; Truong, N. P.; Plebanski, M.; Kedzierska, K.; Mahanty, S.; Chung, A. W.; Caruso, F.; Wheatley, A. K.; Juno, J. A.; Kent, S. J., Anti-PEG Antibodies Boosted in Humans by SARS-CoV-2 Lipid Nanoparticle mRNA Vaccine. *ACS Nano* **2022,** *16,* 11769.

5) Rawstron, A. C.; Villamor, N.; Ritgen, M.; Böttcher, S.; Ghia, P.; Zehnder, J. L.; Lozanski, G.; Colomer, D.; Moreno, C.; Geuna, M.; Evans, P. A. S.; Natkunam, Y.; Coutre, S. E.; Avery, E. D.; Rassenti, L. Z.; Kipps, T. J.; Caligaris-Cappio, F.; Kneba, M.; Byrd, J. C.; Hallek, M. J.; Montserrat, E.; Hillmen, P., International standardized approach for flow cytometric residual disease monitoring in chronic lymphocytic leukaemia. *Leukemia* **2007**, *21,* 956.
